# Deriving LD-adjusted GWAS summary statistics through linkage disequilibrium deconvolution

**DOI:** 10.64898/2026.04.10.26350574

**Authors:** Asma Nouira, Martin Tournaire, Mario Favre Moiron, Marie Verbanck

**Author notes:** These authors contributed equally to this work (Asma Nouira, Martin Tournaire, Mario Favre Moiron). Correspondence: Marie Verbanck < >.

## Abstract

Genome-wide association studies (GWAS) have identified numerous genetic variants associated with complex traits. However, linkage disequilibrium (LD) confounds these associations, leading to false positives where non-causal variants appear associated because they are correlated with nearby causal variants. This is particularly the case in highly polygenic traits where the genome can be saturated in causal variants. To address this issue, we propose *LDeconv* a method based on truncated singular value decomposition (SVD) that adjust GWAS summary statistics without requiring individual-level genotype data. This approach accounts for LD structure, isolates causal variants in high-LD regions, and improve the reliability of effect size estimates. We assessed the performance of *LDeconv* through simulations across various LD scenarios, conducted extensive sensitivity analyses, and applied the method to real GWAS data from the UK Biobank. Our results demonstrate that *LDeconv* effectively reduces false discoveries while preserving true associations, offering a robust framework for post-GWAS analyses, such as Mendelian Randomization.

## Introduction

Genome-wide association studies (GWAS) have provided valuable insights into the genetic basis of complex traits, and led to the identification of over a million unique associations between genetic variants and a wide variety of traits, according to GWAS catalog^1,2^. Despite these advancements, a significant gap remains between the discovery of genetic associations and their translation into biological understanding^3^. Many reported associations lack independent validation, and the precise biological mechanisms connecting these variants to specific traits often remain poorly understood. One of the key challenges in interpreting GWAS findings is linkage disequilibrium (LD): because recombination rarely separates physically close variants, their alleles tend to be inherited together and are therefore correlated. This correlation can confound the interpretation of genetic associations, making it difficult to distinguish between true causal variants and those that are merely correlated with causal ones^4^. Thus, variants in high LD may exhibit inflated effect sizes, which complicates the identification of the true causal variant(s)^5^.

In response to this challenge, several methods have been developed to account for LD using various statistical frameworks. Fine mapping methods, such as FINEMAP^6^, SuSiE^7^, SuSiEx^8^, CARMA^9^ or CAVIAR^10^, aim to pinpoint specific genetic variants within an LD block by estimating the probability of each variant being causal, and typically single out a group of variants whose posterior probabilities of being causal exceed a certain threshold. This is generally accomplished through Bayesian statistical frameworks which integrate prior knowledge, specifically the LD correlation structure, with observed effect sizes. Another popular approach, the COJO method^11^, identifies independent signals within a locus by performing conditional analysis, adjusting the marginal variant effects to account for the LD structure estimated from reference individual-level data. Similarly, LDpred^12^ improves the accuracy of polygenic risk scores (PRS) by adjusting variant effect sizes based on the LD structure. While these methods are powerful, they primarily focus on improving prediction accuracy or identifying independent signals, but do not fully address the challenge of deconvoluting effect sizes resulting from LD correlations. Another proposed approach involves LD-corrected genomic relationship matrices^13^, which uses Mahalanobis distance to adjust for LD in estimating variant-heritability within mixed models. However, they require individual-level genotype data and are designed to model relationships between individuals rather than adjusting marginal variant effect sizes.

Here, we introduce *LDeconv*, a novel approach that directly tackles this issue of deconvoluting GWAS effect sizes in the presence of LD. Using GWAS summary statistics, our method adjusts association signals for LD and aims to isolate the causal variant(s) within loci associated with multiple correlated variants, and provides a deconvolved estimate of the variant effect size. This approach improves our ability to infer causal relationships in GWAS while mitigating false positives induced by LD, making it a more robust tool for understanding the genetic underpinnings of complex traits. The resulting deconvolved summary statistics are particularly suited for post-GWAS analyses.

Our approach operates on summary statistics and directly targets the deconvolution of LD-induced inflation in variant-level association estimates, enabling more interpretable and scalable post-GWAS analysis. One important application of *LDeconv* is Mendelian randomization (MR), a causal inference framework that estimates relationships between traits using genetic variants as instrumental variables (IVs). A major limitation is that LD can inflate marginal variant effect estimates and bias MR results, notably by overestimating causal effects^14,15^. A common mitigation is LD clumping of IVs, *i*.*e*. retaining a set of nearly independent variants, typically by selecting the most significant variant in each LD region^16,17^. However, clumping does not correct the estimated effect sizes for LD-driven inflation from neighboring variants, instead, it merely ensures that correlated signal is not counted multiple times. In contrast, using deconvolved LD-adjusted effect estimates in MR can directly target this source of bias and should therefore yield more accurate causal effect estimates between traits in MR analyses.

## Materials and methods

We introduce *LDeconv*, an LD deconvolution approach for GWAS summary statistics that aims to isolate true causal effects obscured by LD, using Truncated Singular Value Decomposition (TSVD), and to estimate variants deconvolved effect sizes. (Figure 1).

**Figure 1.**
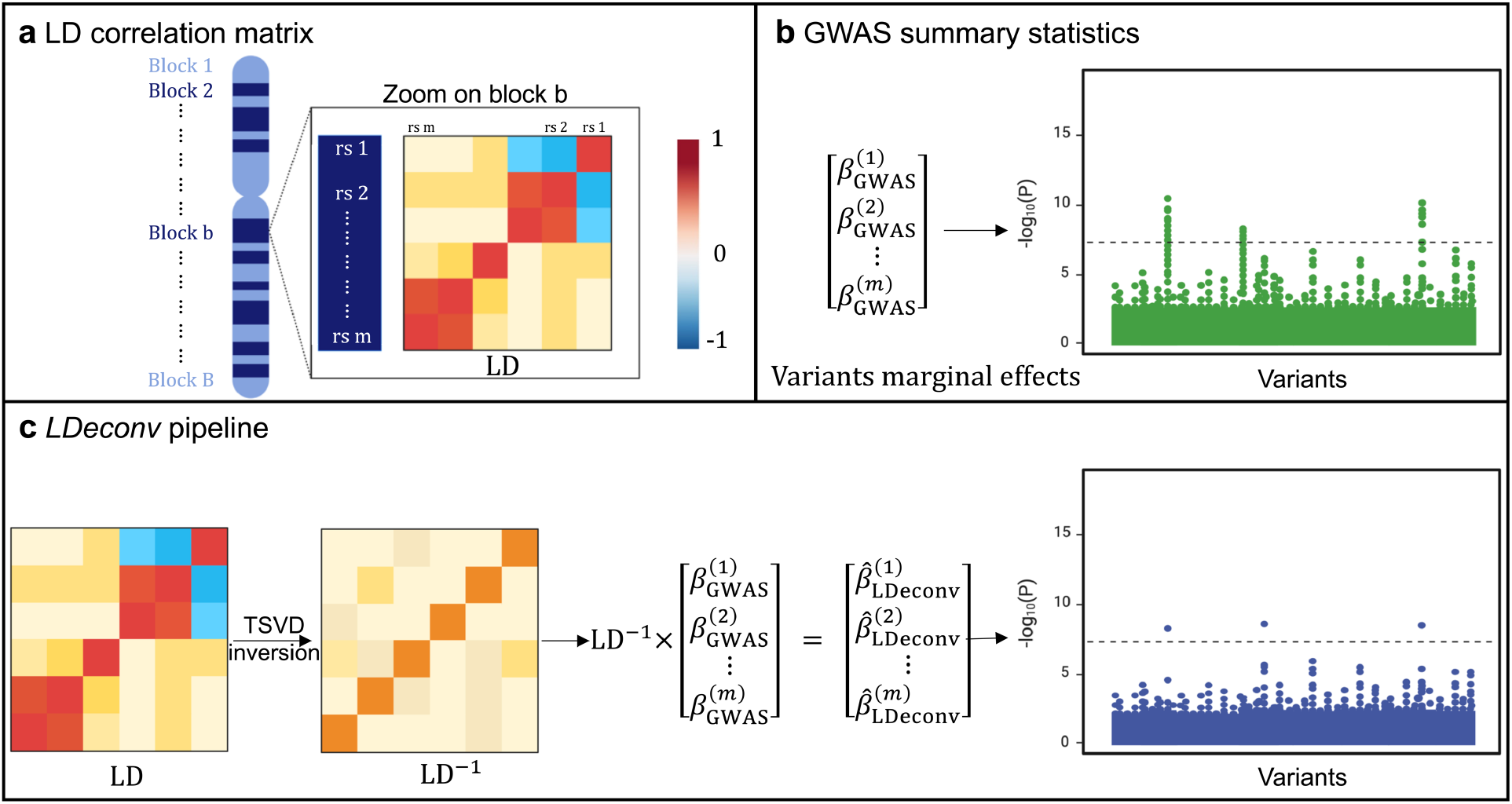
**a** Chromosome-wide linkage disequilibrium (LD) correlation matrix organized into LD blocks, with a zoomed-in view of one representative block showing local correlation structure among genetic variants. **b** GWAS summary statistics represented as a vector of marginal genetic variant effect estimates, and their corresponding association signals across a genomic locus. **c** LDeconv pipeline. For each LD block, the **LD** matrix is inverted using truncated singular value decomposition (TSVD) to obtain a pseudo-inverse **LD**^+^. This pseudo-inverse matrix is then applied to the vector of GWAS marginal effects to compute LD-adjusted effect estimates, thereby reducing LD-induced inflation and sharpening localization of putatively causal variants.

The proposed method operates without requiring individual-level genotype data and is designed to improve the interpretability of GWAS signals by accounting for the correlation between genetic variants due to LD. Specifically, the TSVD-based approach provides a stable matrix decomposition-based inversion of the sparse LD structure, allowing the recovery of adjusted variant-level effect estimates. Below, we present the problem formulation and the proposed method in detail.

### General framework of *LDeconv*

Let *m* denote the number of genetic variants considered within a genomic locus. The LD structure among these *m* variants is represented by the matrix **LD** ∈ ℝ^*m*×*m*^, whose entries quantify pairwise correlation between allele dosages. Specifically, the element **LD**_*ij*_ corresponds to the Pearson correlation coefficient, *r*_*ij*_, between variant *i* and variant *j*, computed as:

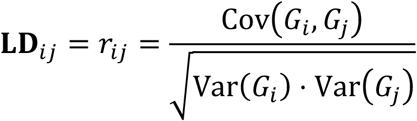

where *G*_*i*_ and *G*_*j*_ denote the allele dosage vectors for variants *i* and *j* across individuals in a population. The matrix **LD** is symmetric, *i*.*e*., **LD**_*ij*_ = **LD**_*ji*_ for all *i, j*, and has unit diagonal elements, ∀*i*, **LD**_*ii*_ = 1, since each variant is perfectly correlated with itself. Variants are assumed to be ordered by physical genomic position so that the correlation structure reflects local LD patterns along the chromosome. Let *β*_GWAS_ ∈ ℝ^*m*^ denote the vector of marginal effect sizes from GWAS, and let *β*_causal_ ∈ ℝ^*m*^ represent the underlying true causal effects of the corresponding variants on the trait.

In GWAS, observed marginal effect sizes *β*_GWAS_ are confounded by LD because the estimated effect of each variant captures not only its own contribution *β*_causal_ to the trait, but also the contributions of nearby correlated variants. This relationship can be expressed as:

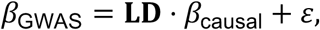

where ε represents the error term and **LD** represents the LD correlation matrix.

To accurately estimate the true causal effects *β*_causal_, we propose to multiply the GWAS effect sizes *β*_GWAS_ by the inverse of the LD matrix. The causal effects could then be estimated as:

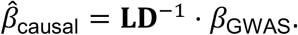

For any variant *j*, 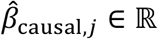 represents its estimated causal effect on the trait.

However, in practice, the LD matrix is often not invertible or numerically unstable. Algebraically, a square matrix **LD** ∈ ℝ^*m*×*m*^ is invertible if and only if it is full rank, meaning that rank(**LD**) = *m*. This condition requires all singular values of the matrix **LD** to be non-zero and its columns (and rows) to be linearly independent.

In real genomic data, these assumptions are likely violated because nearby variants exhibit strong correlations resulting from shared recombination history. Hence, many variants can be approximately represented as linear combinations of neighboring variants, leading to multicollinearity and rank deficiency. As a result, the LD matrix often contains extremely small or zero singular values, making direct inversion either undefined or numerically unstable.

To overcome this, we propose to use the Moore-Penrose pseudo-inverse, denoted **LD**^+^, which generalizes matrix inversion to singular or ill-conditioned matrices. The pseudo-inverse satisfies the following four properties:

1. **LD LD**^+^ **LD** = **LD**

2. **LD**^+^**LD LD**^+^ = **LD**^+^

3. (**LD LD**^+^)^⊤^ = **LD LD**^+^

4. (**LD**^+^**LD**) ^⊤^ = **LD**^+^ **LD**

When **LD** is full rank, the pseudo-inverse coincides with the true inverse: **LD**^+^= **LD**^−^. Moreover, to improve numerical stability, we propose to apply truncated singular value decomposition (TSVD), retaining only the top *k* singular values for pseudo-inverse construction. This helps reduce the contribution of very small singular values, which are often dominated by noise, while keeping the main LD patterns required for effect-size deconvolution. Importantly, we explored several methods to calibrate the strategy to estimate the LD pseudo-inverse matrix, and chose to use TVSD which was the best compromise based on simulations (see Supplementary Results, Supplementary Figures 1 and 2).

**Figure 2.**
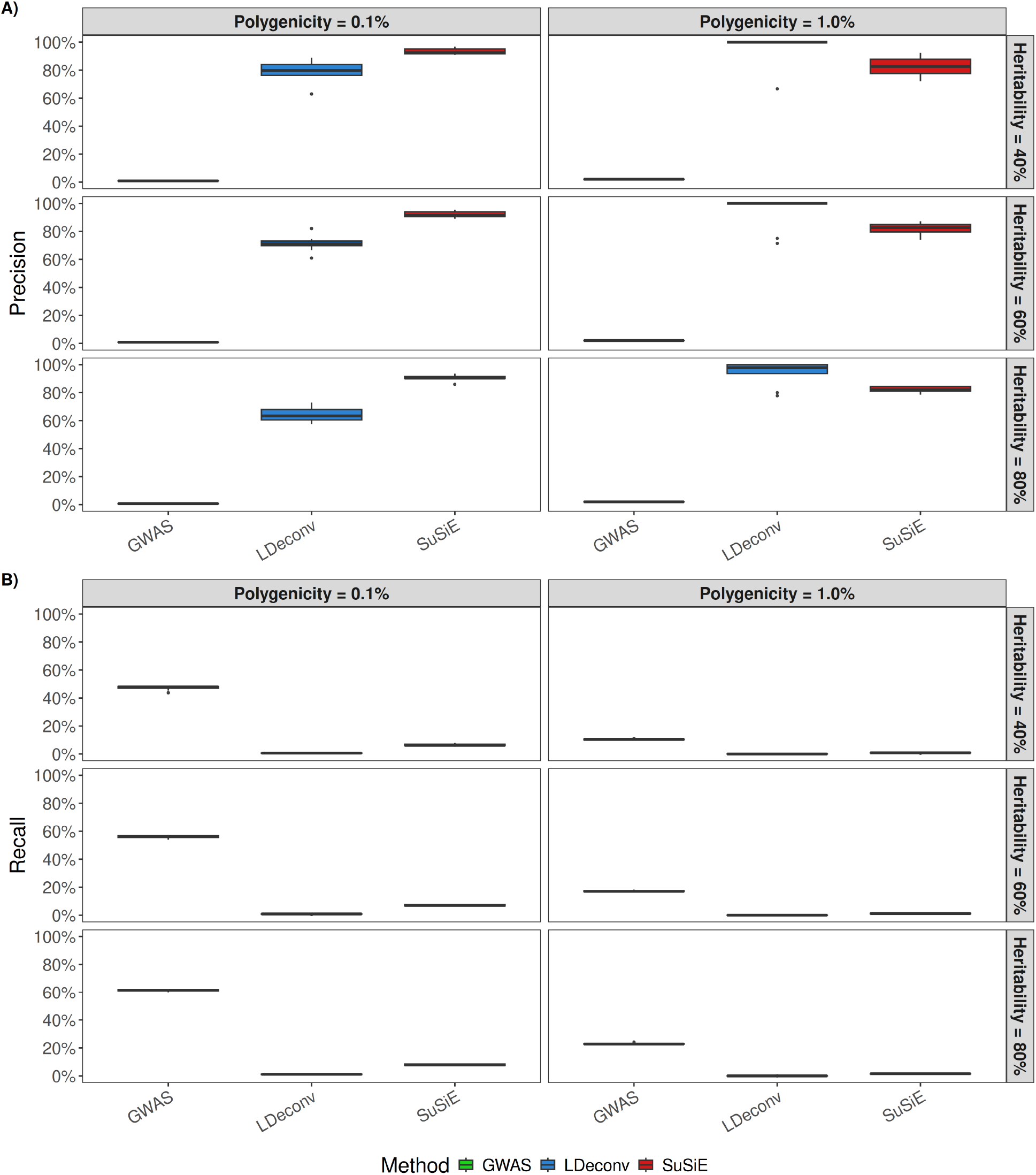
A) Precision and B) Recall of predicted causal variants in simulations. The x-axis shows the three methods used to identify causal variants, colored by method: green for the observed GWAS results, blue for LDeconv, and red for SuSiE. For a given genetic variant, statistical significance was defined as p < 5 × 10^−8^ for GWAS and LDeconv, and as PIP > 0.9 for SuSiE. Results are shown across six simulation scenarios (panels) spanning combinations of polygenicity in columns and heritability in rows. Here, polygenicity denotes the proportion of independent variants that are truly causal to the simulated trait, and heritability is the proportion of trait variance explained by genetic effects. Each boxplot summarizes the performance over 10 independent replicates. Simulated sample size is 500,000.

### *LDeconv*: Truncated Singular Value Decomposition for LD Deconvolution

#### Local approximation of the LD matrix by sliding windows

To recover true causal effect sizes from GWAS summary statistics confounded by linkage disequilibrium (LD), we use a deconvolution method based on truncated singular value decomposition (TSVD) applied to local approximations of the LD matrix. The chromosome-wide LD structure is highly localized: variants within sufficiently small genomic windows tend to exhibit strong correlations, while LD rapidly decays with distance. Hence, the full LD matrix can be well-approximated by an arrangement of local blocks.

Therefore, we partition the LD matrix **LD** ∈ ℝ^*m*×*m*^ into *B* non-overlapping blocks such that:

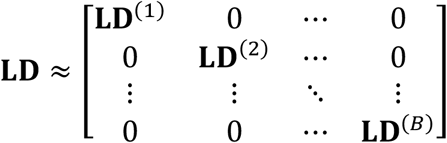

where each 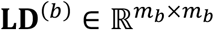 is the local LD matrix for block *b*, with 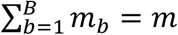. For computational reasons, we constrain block sizes to *m*_3_ < 10,000. To minimize the loss of LD at block boundaries under this constraint, we use the `AdjClust` R package^18,19^ to define adjacency-constrained blocks that best respect local LD. Concretely, for each chromosome, we first construct strictly independent blocks using an igraph-based approach. Second, for any independent block exceeding the size constraint, we perform hierarchical clustering within the block using `adjClust(h=10000)` on the corresponding LD submatrix, and use `cutree_chac()` to select the minimum number of clusters such that all resulting clusters satisfy *m*_3_ < 10,000. The resulting adjacency-constrained clusters define our final LD blocks.

Because *LD* is not strictly block-diagonal, inverting each block separately may ignore residual correlations between neighboring regions and introduce boundary artifacts. Thus, to better capture local LD structure, we use overlapping sliding windows composed of three consecutive LD blocks. For each block *b* = 2, …, *B* − 1, we form a local 3 × 3 block LD matrix containing the block itself together with its immediate neighboring blocks (*b* − 1, *b, b* + 1):

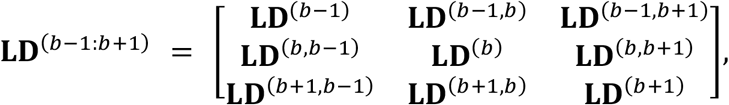

where **LD**^(*i,j*)^ denotes the LD submatrix between blocks *i* and *j*. The diagonal terms correspond to within-block LD matrices, and the off-diagonal terms represent LD correlations between neighboring blocks.

### TSVD-based pseudo-inverse

Within each sliding window, we perform truncated SVD, retaining the top *k* singular values corresponding to 99% of the variance. The corresponding truncated pseudo-inverse is:

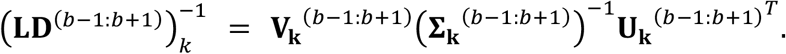

where **U**_**k**_ and **V**_**k**_ denote the matrices of the retained left and right singular vectors, respectively, and **Σ**_**k**_ is the diagonal matrix containing the retained singular values.

### Deconvolution of GWAS summary statistics

We then apply this three-window-wise pseudo-inverse 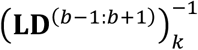 to the vector of GWAS estimates framing block 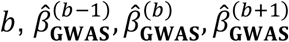, which results in:

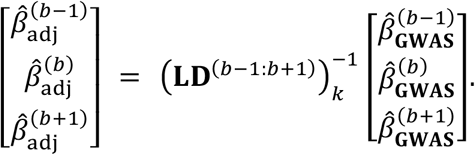

Finally, we only keep the block from the middle to build our estimator of the variant causal effects 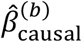 :

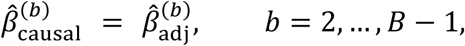

and construct the genome-wide adjusted effect sizes by concatenation of the block-wise estimates 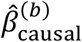 :

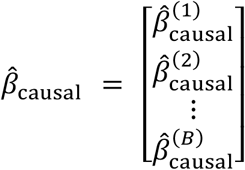

For the first and last blocks, we apply the same strategy using the available neighboring blocks (*e*.*g*. (1: 2) and (*B* − 1: *B*)), while still retaining only the block-specific component for the estimation. This approach aligns with the known genomic structure of LD into blocks and offers a practical alternative to inverting the full LD matrix, which is often computationally expensive and numerically unstable. Using these smaller blocks, the method remains efficient while still capturing local LD patterns. That being said, since the LD matrix is not strictly block-diagonal, defining block boundaries requires setting a threshold, a step that involves balancing between computational simplicity and the accuracy of the LD representation. We therefore propose a quality score to identify variants for which a substantial part of the LD is outside the window used for deconvolution. Briefly, we defined the quality score for each variant as the proportion of its chromosome-wide LD score captured within the three-block-window used for deconvolution (Supplementary Results, Supplementary Figures 3 and 4). Finally, we verify that the resulting inverse matrices satisfy the Moore-Penrose properties (see Supplementary Results and Supplementary Table 1).

**Figure 3.**
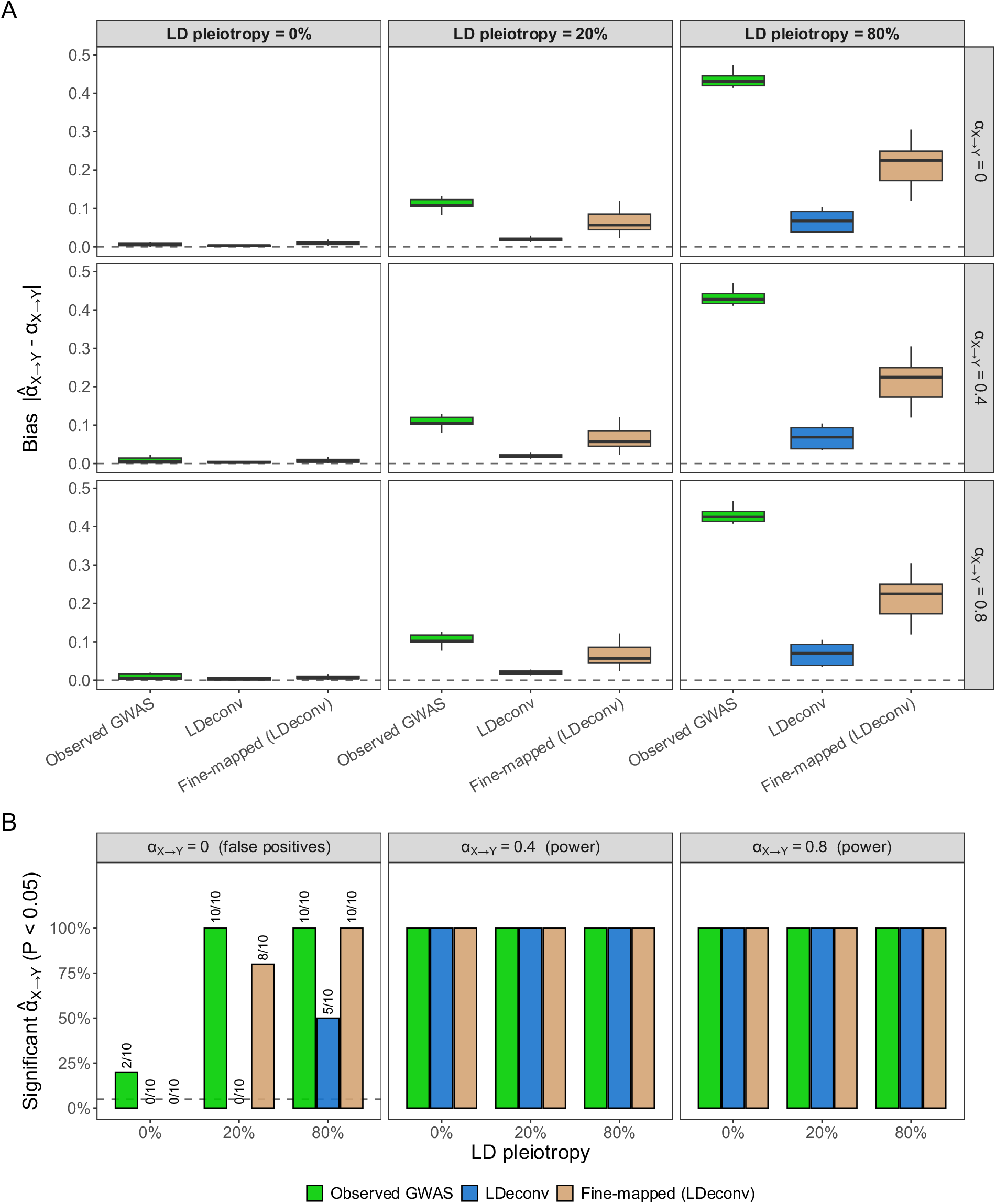
Mendelian Randomization under directional linkage disequilibrium (LD) confounding. A) Absolute bias of the IVW causal effect estimate, 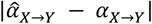, for the three MR strategies across a gradient of directional LD pleiotropy (0% to 80%, columns) and of simulated causal effects α_X→&_ (rows). B) Proportion of replicates with a significant IVW causal effect estimate (P < 0.05). Under the null hypothesis (H_0_: α_X→&_ = 0), this proportion corresponds to the false-positive rate (dashed line: nominal 5% level); under the alternative hypothesis (H_1_: α_X→&_ ≠ 0), it corresponds to the statistical power. Observed MR used marginal GWAS summary statistics. Fine-mapped (LDeconv) MR utilized LDeconv-selected variants with marginal effect sizes. Deconvolved MR employs both LDeconv-selected variants and adjusted effect sizes. A standard genetic architecture parameters (N = 500,000; h^2^ = 0.5; polygenicity = 0.001) was simulated.

**Figure 4.**
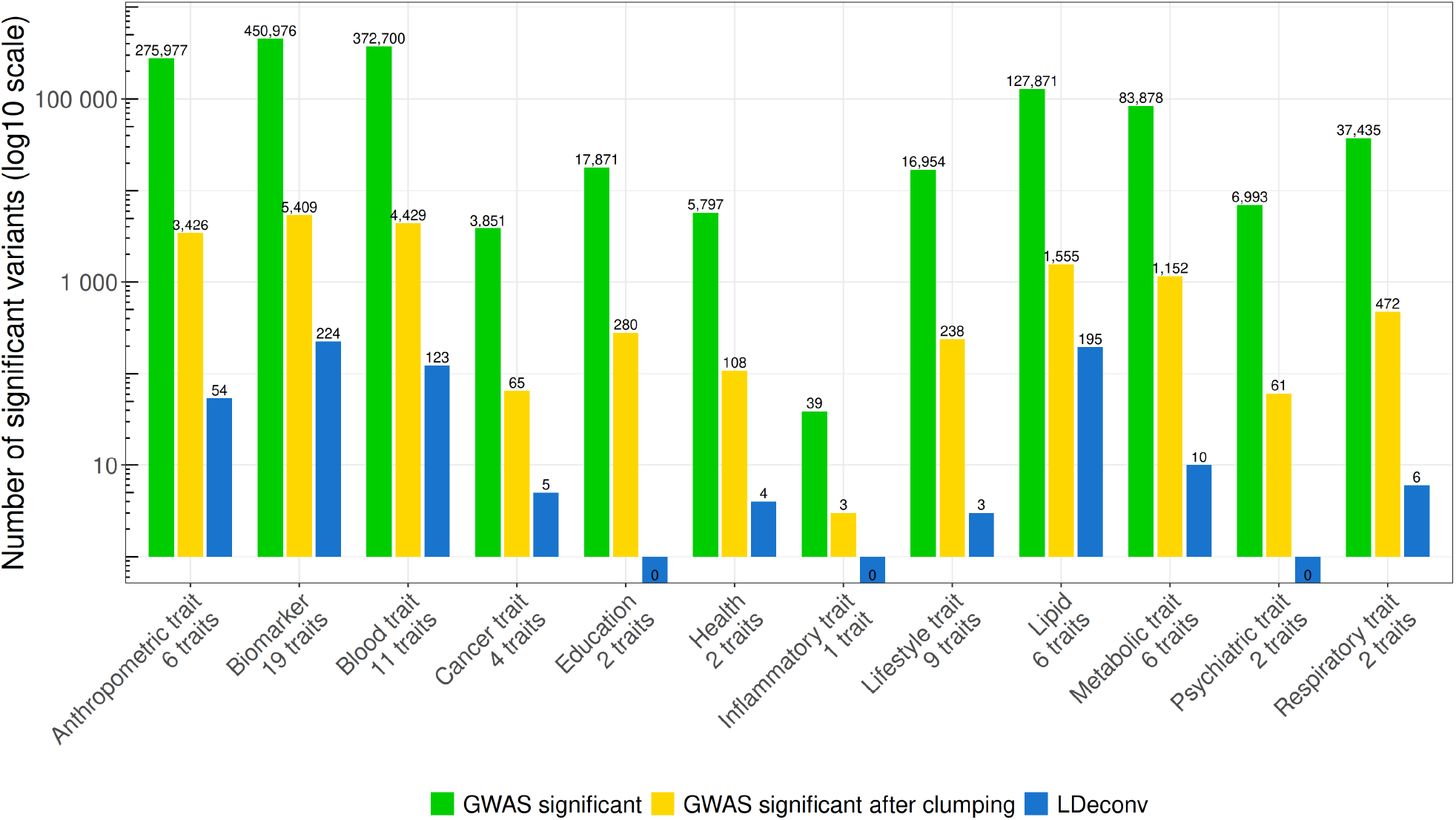
Number of significant variants across categories for 3 different methods. The x-axis shows the phenome categories, and the y-axis indicates the number of significant variants on a log_10_ scale. Bars are grouped by phenome categories and colored as follows: green for GWAS-significant variants, yellow for GWAS-significant variants after clumping with a stringent r^2^ = 0.01 threshold, and blue for LDeconv-significant variants. Statistical significance was defined using the method-specific thresholds described in the Methods section. The number of significant variants in each category is indicated above the bar.

### Deconvolved effect sizes and standard errors

In practice, *LDeconv* was applied to the standardized test statistics, 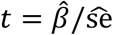, rather than to the marginal effect sizes directly, since a standardized effect measure is more appropriate for comparisons across traits. Deconvolved P-values were derived from the deconvolved *t*-statistics. For post-GWAS analyses that require effect sizes and their standard errors, such as Mendelian randomization, we reconstructed deconvolved effect sizes and standard errors from the deconvolved *t*-statistics similarly previous work^20,21^. For a given variant with deconvolved *t*-statistic *t*, sample size *n*, and minor allele frequency *MAF*, we computed:

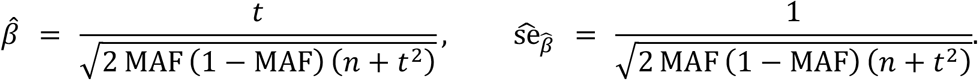

These reconstructed quantities were used in both the simulated and real-data Mendelian randomization analyses.

### Simulation study

To evaluate *LDeconv* under controlled conditions, we designed a simulation framework that modeled the relationships between causal effect sizes, LD structure, and observed GWAS summary statistics in three consecutive steps: 1) generating true causal effects, 2) propagating these effects through LD, and 3) adding observational noise.

### Generating true causal effect

To reflect “realistic” GWAS data and evaluate our method across different genetic architectures, we simulated genetic effects across all 22 autosomes by explicitly modeling an exposure (X), an outcome (Y), and a genetic confounder (U). We denote *m* ~ 3.8 million as the total number of genome-wide variants. The polygenicity of the simulated traits was defined as the expected fraction of causal variants, denoted *λ* ∈ {0.1%, 1%} drawing the exact number of causal variants per chromosome, denoted λ ∈ 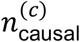, from a binomial distribution, 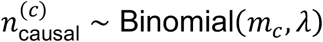, where *m*_*c*_ is the number of variants on the specific chromosome. Causal variants were assigned baseline effect sizes drawn from a normal distribution:

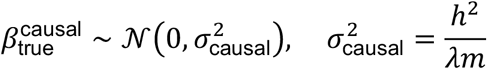

This ensures that the total expected genetic variance matched the input total variant-based heritability *h*^2^ ∈ {0.4,0.6,0.8}. To account for chromosome size differences, the global heritability was weighted according to the proportion of genetic variants on each chromosome. True non-causal variant effects were assigned to zero, *i*.*e*.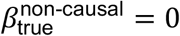.

Importantly, to explicitly evaluate the robustness of *LDeconv* and downstream Mendelian randomization analyses against LD-driven confounding, we introduced a mechanism of directional LD pleiotropy. Rather than randomly selecting variants causal to the outcome, we forced a proportion of variants (0%, 20%, and 80%) to be in strong LD with variants causal to the exposure, with 0% corresponding to the baseline scenario. Concretely, for each exposure causal variant, we selected the neighboring variant with the highest absolute LD correlation (*r*) and assigned it as a causal variant for the outcome, while matching the direction of effect to the LD sign. These simulations were designed to create scenarios where LD could generate false-positive association patterns and inflate downstream causal estimates.

### LD propagation

These true causal effects were then propagated through the LD matrix to generate marginal GWAS signals:

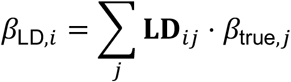

The LD matrices were derived from UK Biobank genotypes for the 22 autosomes, ensuring realistic correlation structures representative of the European population.

### Adding observational noise

To reflect the scale of modern large biobanks and examine the effect of increased statistical power, observational noise was added to simulate three finite sample sizes *N* ∈ {100,000, 500,000, 1,000,000}, producing the observed summary statistics:

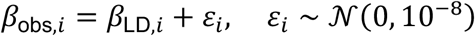

Finally, the standard errors (se) of the observed effect sizes were computed for every variant as: 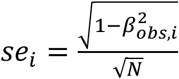.

To account for variability and ensure robustness, we generated 10 independent genome-wide replicates for each combination of polygenicity *λ*, heritability *h*^2^, and sample size *N*, resulting in a comprehensive simulation grid of 18 scenarios (Supplementary Table 2).

### Evaluation Metrics

To evaluate the performances of *LDeconv*, we compared the GWAS-observed marginal effect sizes with those estimated using *LDeconv*. The evaluation was based on two metrics: 1) Precision to quantify the proportion of correctly identified causal genetic variants among the selected significant variants; 2) Recall to quantify the fraction of true causal variants successfully recovered. For each method, significant variants were selected using the conventional genome-wide significance threshold of *P* < 5 × 10^−8^ both in the original and deconvolved summary statistics. Evaluation metrics were computed against the set of known causal variants. This approach reflects how variants would typically be prioritized in real GWAS analyses and ensures comparability across methods. Together, these metrics evaluate the ability of each method to recover true causal signals in the presence of LD.

### Comparison with fine-mapping

To evaluate the ability of *LDeconv* to identify causal effects, we compared our approach with the widely-used fine-mapping method, SuSiE^22^. Importantly, the two methods differ in their outputs: our approach primarily produces LD-adjusted summary statistics, whereas SuSiE outputs credible sets together with variant-level posterior inclusion probabilities (PIP) for regions of interest. We computed PIPs within each region of interest using `susie_rss(L=10, coverage = 0.95, estimate_prior_variance=TRUE, estimate_residual_variance=TRUE)`. Regions of interest were defined following the procedure described by the authors^23^. Briefly, we defined an initial fine-mapping region as all variants within ±250 kb of a significant association. Overlapping regions were then iteratively merged until no overlap remained. For comparison purposes, we considered a variant as “causal” under SuSiE if its PIP > 0.9, as proposed by the authors. Both in simulations and real data, we used GWAS significance (*p* < 5 × 10^−8^) to define the fine-mapped regions, since the objective was to compare the performances for identifying causal variants between *LDeconv* and GWAS.

### Mendelian randomization in simulations

To assess whether LD-adjusted summary statistics could improve downstream causal inference, we performed Mendelian randomization (MR) analyses in an additional set of simulations under a standard genetic architecture (*N* = 500,000, *h*^2^ = 0.5, *λ* = 0.1%) combining a sample size correpsponding to the UK-Biobank scale, a moderate heritability, and the lowest polygenicity, for which *LDeconv* achieved its highest precision. The true effects of the exposure and the outcome were defined as:

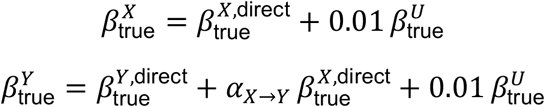

where *α*_*X*→*Y*_ ∈ {0,0.4,0.8} is the true causal effect of ***X*** on *Y* and *U* is the genetic confounder. Each causal effect was combined with three levels of directional LD pleiotropy (0%, 20% and 80% of the outcome causal variants, see above), resulting in nine scenarios, each simulated in 10 independent replicates.

For each scenario, we compared three MR strategies: (i) *Observed* MR, where instrumental variables (IVs) were selected among GWAS-significant exposure variants (*P* < 5 × 10^−8^) and marginal GWAS effect sizes were used for both the exposure and the outcome; (ii) *Fine-mapped (LDeconv)* MR, where IVs were selected among *LDeconv*-significant exposure variants (*P* < 5 × 10^−8^), but marginal GWAS effect sizes were used; and (iii) *Deconvolved* MR, where the IVs of (ii) were used together with deconvolved effect sizes and standard errors (see Methods). In all three strategies, candidate IVs were LD-clumped with PLINK as implemented in the `genal` Python library^24^ (*r*^1^ < 0.01 within a 10,000 kb window, 1000 Genomes European reference panel, GRCh37), retaining in each clump the variant with the smallest exposure P-value (GWAS P-values in (i), deconvolved P-values in (ii) and (iii), which therefore share the same IVs). Causal effects were estimated with the inverse-variance weighted (IVW) method^14^, and performance was assessed across replicates by the absolute bias 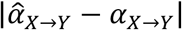 and by the proportion of significant IVW causal estimates (*P* < 0.05), *i*.*e*. the false-positive rate under H_0_(*α*_F→H_ = 0) and the power under H_&_ (*α*_F→H_ ≠ 0).

### Real Data: UK Biobank Summary Statistics

We applied *LDeconv* to real GWAS summary statistics from the UK Biobank^25^, a large-scale population cohort with deep phenotyping and genotype data. We first selected a set of 70 representative heritable traits spanning 12 broad phenome categories (Supplementary Table 3). We chose to particularly focus on lipid metabolism and metabolic diseases, known to exhibit polygenic architectures and LD-driven pleiotropy. For Mendelian randomization, we selected known exposure-outcome pairs to act as positive and negative controls (Supplementary Tables 3 and 4).

For each trait, we used publicly available GWAS summary statistics computed on individuals of European ancestry. Sex chromosomes were excluded due to their specific inheritance patterns and sex-dependent biology. We further excluded low-confidence variants and rare variants with minor allele frequency below 0.05, which have limited statistical power, as well as the few variants with over 2 alleles, for which LD handling is not straightforward in our framework. LD matrices for the retained variants were reconstructed from the full LD matrices provided by UK Biobank. This came down to consider ~3.8 million variants, both for the real data analyses and the simulations. When specified, clumping was performed for using the `ieugwasr::ld_clump()` R function^26^. These data allowed us to evaluate the relevance of *LDeconv* in real-world settings where the LD structure and signal sparsity are both complex and trait-specific.

### Biological Interpretation of Real Data

Beyond simulation-based evaluations, we investigated the biological relevance of our results obtained from real UK Biobank summary statistics. Specifically, we examined how *LDeconv* reshaped association patterns within regions containing dense GWAS signals and evaluated the biological plausibility of recovered variants through several layers of downstream analysis.

We performed LocusZoom visualizations for selected genomic regions harboring a high density of significant associations. These regions were chosen based on genome-wide significance (GWAS threshold *P* < 5 × 10^−8^) and high local LD. By comparing GWAS signals with the outputs of *LDeconv*, we examined how *LDeconv* reduced the relative prominence of variants within a locus and whether the deconvolved signals offered improved resolution of putative causal variants.

### Mendelian randomization in real data

To illustrate the practical interest of the adjusted summary statistics produced by *LDeconv*, we performed Mendelian randomization (MR) analyses using, on one hand, the original UK Biobank GWAS summary statistics, and on the other hand, the *LDeconv* summary statistics. Importantly, we used the same set of instrumental variables (IVs) for both methods: IVs were defined as variants reaching significance according to both GWAS and *LDeconv*, so that any difference in MR results was attributable to the estimated summary statistics used rather than to IV selection.To compare MR performance between GWAS summary statistics and *LDeconv* summary statistics, we adopted a positive/negative control framework^20^. We selected pairs of UK Biobank traits as positive controls (established biological causal effects) and negative controls (no expected causal effect) (see Figure 6 and Supplementary Table 4). Positive controls mainly consisted of established biomarker-disease effects. Negative controls were based on a systematic test of adult hair color on childhood traits (birth weight, body size at age 10, and maternal smoking at birth), for which no causal effect is expected. MR was then performed using the inverse-variance weighted (IVW) method^14^ for exposures with ≥ 3 IVs, or the Wald ratio otherwise. We also computed the *I*^2^ index^27,28^, which quantifies the heterogeneity of the individual IV Wald estimates.

For the following set of P-value thresholds {0, 10^−5^, 10^−4^, 10^−3^, 10^−2^, 0.05,0.1,0.5,1}, we evaluated MR performances by computing the precision and recall as a function of the significance threshold applied to the MR test. Precision was computed using only negative controls, as the proportion of correctly predicted non-significant effects across negative-control pairs. Recall was computed using only positive controls, as the proportion of correctly predicted significant effects across positive-control pairs. Varying the significance threshold allowed us to obtain a ROC curve (Supplementary Figure 5) and compute the corresponding area under the curve (AUC) as a global performance metric.

**Figure 5.**
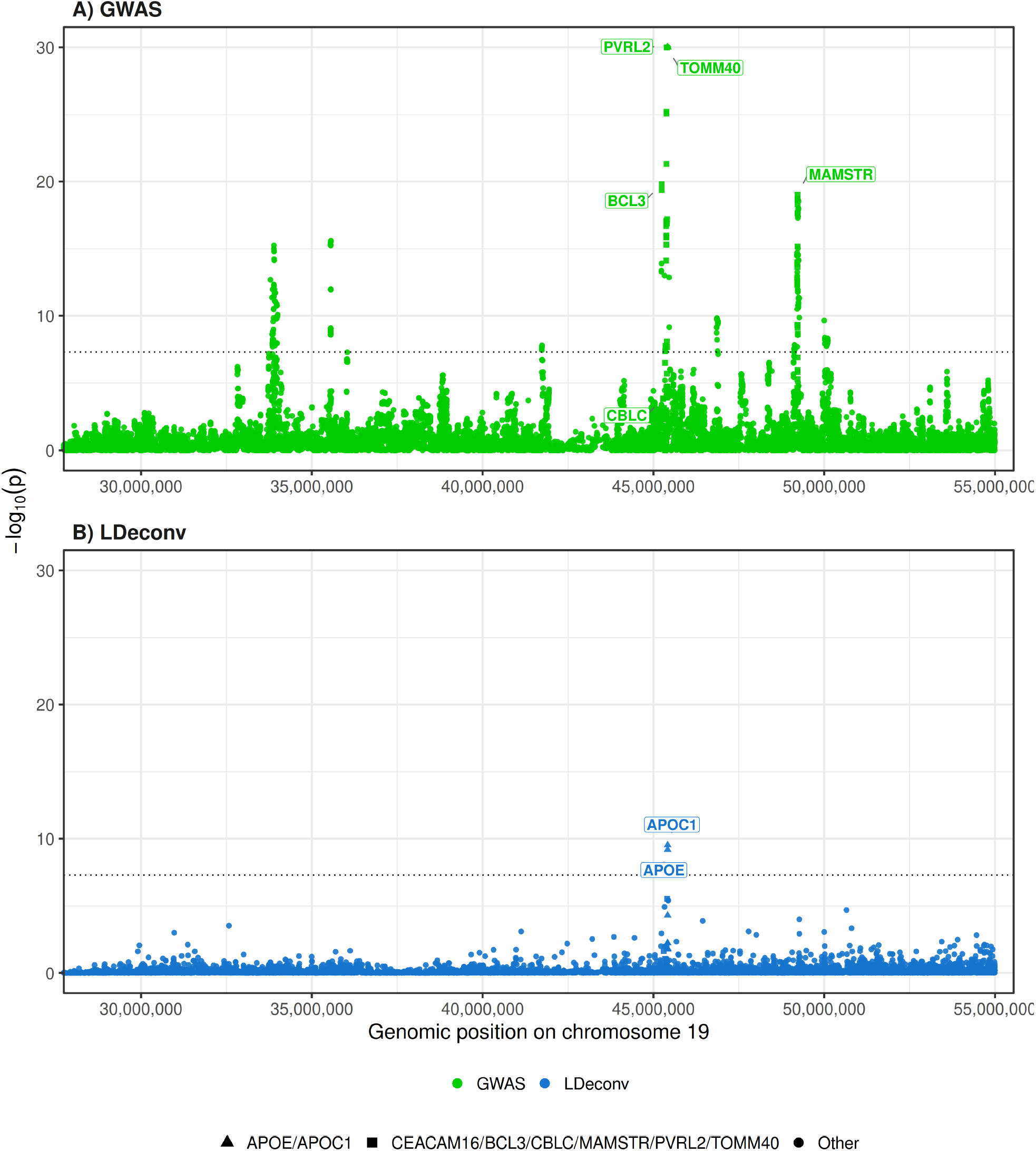
Locus plot for triglyceride levels on chromosome 19, for A) GWAS and B) LDeconv. The x-axis spans genomic positions 25,000,000-55,000,000. The y-axis shows the –log_10_ association P-values for individual variants, colored by method: green for the observed GWAS results and blue for LDeconv. Variants annotated to APOE and APOC1 are represented as triangles. Variants annotated to CEACAM16, BCL3, CBLC, PVRL2, and TOMM40 are represented as squares. All other variants are represented as circles. The dotted line corresponds to the significance threshold at p < 5 × 10^−8^. All P-values < 10^−30^ were set at 10^−30^ for readability.

## Results

### *LDeconv* reduces false discoveries by controlling LD-induced inflation in simulations

Through our simulation study, we evaluated *LDeconv* across different genetic architectures by varying polygenicity, heritability, and sample size (See Methods and Supplementary Table 2). We found that *LDeconv* effectively reduces the inflation of marginal GWAS effect sizes caused by local LD. As shown in Figure 2 (N = 500,000), *LDeconv* achieved higher precision than the observed GWAS summary statistics. In low to moderate polygenicity scenarios (0.1% and 1%), *LDeconv* maintained precision between 60% and 100%. In contrast, the precision of standard GWAS remained close to zero due to pervasive LD confounding. This demonstrates that when the genetic architecture is relatively sparse, *LDeconv* is effective at isolating the exact causal variants.

We observed that polygenicity is the primary parameter driving *LDeconv* performance. Higher polygenicity results in smaller individual causal effect under fixed total per-variant heritability, thus in denser genetic landscapes, true signals become harder to distinguish from correlated background signal. Furthermore, statistical power, modulated jointly by sample size and heritability, played a critical role in causal variant discovery. In low-power scenarios, such as *N* = 100,000 (Supplementary Figure 6), the true effect sizes were often too weak to overcome observational noise. This resulted in a lack of genome-wide significant variants passing the *P* < 5 × 10^−8^ threshold. Conversely, increasing the sample size to 500,000 (Figure 2) and 1,000,000 (Supplementary Figure 7), or increasing the overall heritability, effectively restored statistical power. Higher power improves the signal-to-noise ratio, allowing the TSVD to better capture the dominant variance components associated with true effects and to extract causal variants. Our sensitivity analyses showed that *LDeconv* is robust to minor LD discrepancies, but its precision depends on an ancestrally matched LD reference (Supplementary Results, Supplementary Figures 8 and 9).

While standard GWAS identified a higher fraction of true causal variants, it yielded a high rate of false positives. Conversely, *LDeconv* increased precision at the expense of recall, meaning that although *LDeconv* may have missed some true associations, its reduction of false discoveries provided more reliable effect estimates. To further contextualize our performance, we included a comparison with the prominent fine-mapping method SuSiE (Figure 2). Overall, *LDeconv* showed performance comparable to SuSiE across genetic architectures. While SuSiE maintained slightly higher recall and showed better precision at lower polygenicity (0.1%), *LDeconv* demonstrated superior precision under higher polygenicity conditions (1%). Importantly, however, the two methods have distinct aims and outputs. SuSiE generates credible sets and variant-level posterior inclusion probabilities (PIPs), but does not provide adjusted effect sizes. In contrast, *LDeconv* explicitly deconvolves the summary statistics to output corrected marginal effect estimates. As a result, our method is particularly well-suited for post-GWAS applications such as Mendelian randomization, which require effect sizes that closely approximate the truth, combined with a high-confidence set of associated variants.

### *LDeconv* provides Mendelian Randomization estimates that are more robust to directional LD pleiotropy

Standard MR assumes that instrumental variables (IVs) influence the outcome only through the exposure; however, this exclusion restriction assumption is frequently violated when an outcome causal variant is in linkage disequilibrium (LD) with the exposure IV. To quantify the ability of *LDeconv* to mitigate this bias, in simulations, we compared three IV selection procedures: (i) Observed MR, using GWAS threshold to select IVs and marginal effect sizes in the MR model; (ii) Fine-mapped (*LDeconv*) MR, using *LDeconv* to select IVs but marginal effect sizes in the MR model; and (iii) Deconvolved MR, using both *LDeconv*-selected IVs and deconvolved effect sizes. The IVs between ii) and iii) are therefore the same but the effect sizes used in the MR model are different. We evaluated the performance of the three MR models under a standard genetic architecture (N = 500,000; h^2^ = 0.5; polygenicity = 0.001) across a gradient of directional LD confounding (See Methods). Bias was defined as the deviation of the Inverse-Variance Weighted (IVW) estimate from the true simulated effect 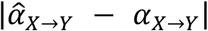. As illustrated in Figure 5A, all three MR models yielded unbiased estimates in the absence of directional LD confounding (median absolute bias ≤ 0.01). However, a clear divergence emerged as the proportion of pleiotropic variants increased. This trend was consistent across both the null (H_0_: *α*_*X*→*Y*_ = 0) and alternate hypothesis (H_1_: *α*_*X* →*Y*_ ≠ 0) scenarios. Specifically, at the maximum confounding level (LD pleiotropy = 80%) under H_0_, the bias in the Observed MR design reached a median absolute deviation of 0.43. The Fine-mapped (*LDeconv*) approach (selecting variants via deconvolved P-values but using marginal effect sizes) only partially mitigated this inflation, with a residual median bias of 0.23. This confirms that selecting the causal variant (fine-mapping) was insufficient to ensure the validity of MR if the effect size estimates remain contaminated by correlated signals from LD proxies. In contrast, *LDeconv* effectively decoupled the causal signal from the local LD structure. Specifically, at the maximum confounding level, the resulting IVW estimates maintained a residual median absolute deviation of 0.07. This limited increase across the pleiotropic gradient demonstrated that *LDeconv* effectively accounts for the LD-driven correlation between exposure and outcome signals. However, a biased causal estimate is only problematic in practice if it leads to the spurious detection of a causal effect. We therefore also assessed the false-positive rate, defined as the proportion of replicates in which a significant causal effect (*P* < 0.05) was detected under **H**_0_ (Figure 5B). In the absence of directional LD pleiotropy, the false-positive rate remained low for all three MR models (2/10 replicates for Observed MR and 0/10 for both *LDeconv*-based approaches). With 20% of pleiotropic variants, Observed MR detected a spurious causal effect in all replicates (10/10) and Fine-mapped (*LDeconv*) MR in 8/10 replicates, whereas Deconvolved MR did not yield any false positive (0/10). At the maximum confounding level, Observed and Fine-mapped (*LDeconv*) MR were significant in all replicates, while Deconvolved MR halved the false-positive rate (5/10). Under **H**_1_, all three MR models detected the causal effect in every replicate (10/10), indicating that the reduction of false positives achieved by *LDeconv* did not come at the cost of statistical power. By adjusting the effect sizes themselves, *LDeconv* substantially reduced both the bias and the false positives induced by directional LD pleiotropy, improving the reliability of causal inferences even in genomic regions saturated with correlated signals, although residual false positives persisted under extreme LD pleiotropy.

### *LDeconv* reduces LD-induced signal inflation in real data

We applied *LDeconv* to UK Biobank GWAS summary statistics for 70 heritable traits spanning the phenome and grouped into 12 broad categories. In Figure 4 and Supplementary Table 5, for each trait and phenome category, we report the number of significant associations detected by GWAS, after LD-clumping, and after *LDeconv*. Interestingly, biomarkers and lipid traits retained more significant associations than other categories, possibly reflecting a stronger biological signal. Conversely, Education and Psychiatric traits did not show any significant *LDeconv* associations. These results suggest that many GWAS-significant variants for complex traits may correspond to correlated association signals rather than independent causal effects.

### *LDeconv* prioritizes known variants in Triglycerides on real data

As an illustrative application, we applied *LDeconv* to UK Biobank GWAS summary statistics for triglyceride levels (Figure 5). We then compared the genes harboring the top variants between the raw GWAS associations and the deconvolved associations. Interestingly, variants identified by *LDeconv* were located in genes APOE (Apolipoprotein E) and APOC1 (Apolipoprotein C-I) that are confirmed to be involved in triglyceride metabolism based on previous studies^29–31^. Indeed, APOE regulates the uptake of triglyceride-rich lipoproteins via receptor-mediated pathways, while APOC1 modulates this process by inhibiting lipoprotein lipase and the binding of lipoproteins to hepatic receptors, leading to changes in circulating triglyceride levels. Conversely, the top significant variants identified by GWAS mapped to genes CEACAM16, BCL3, and CBLC which are not known to play a major role in triglyceride metabolism. Together, these results illustrate how *LDeconv* can sharpen GWAS results and help refine association signals toward likely causal variants from summary statistics. It is worth mentioning that SuSiE did not prioritize any variant in this region, despite being more powerful genome-wide than *LDeconv*.

### *LDeconv* summary statistics improve Mendelian randomization robustness

Mendelian randomization (MR) allows to estimate and test the causal effect of an exposure on an outcome using genetic variants as instrumental variables (IVs). As shown in simulations, because LD can inflate marginal GWAS effects, LD-induced pleiotropy can propagate to MR and increase false positives, as well as bias causal estimates. Using 14 positive-control and 19 negative-control exposure-outcome pairs in real data (see Methods), we compared IVW results obtained with *LDeconv* versus original GWAS estimates (Figure 6).

**Figure 6.**
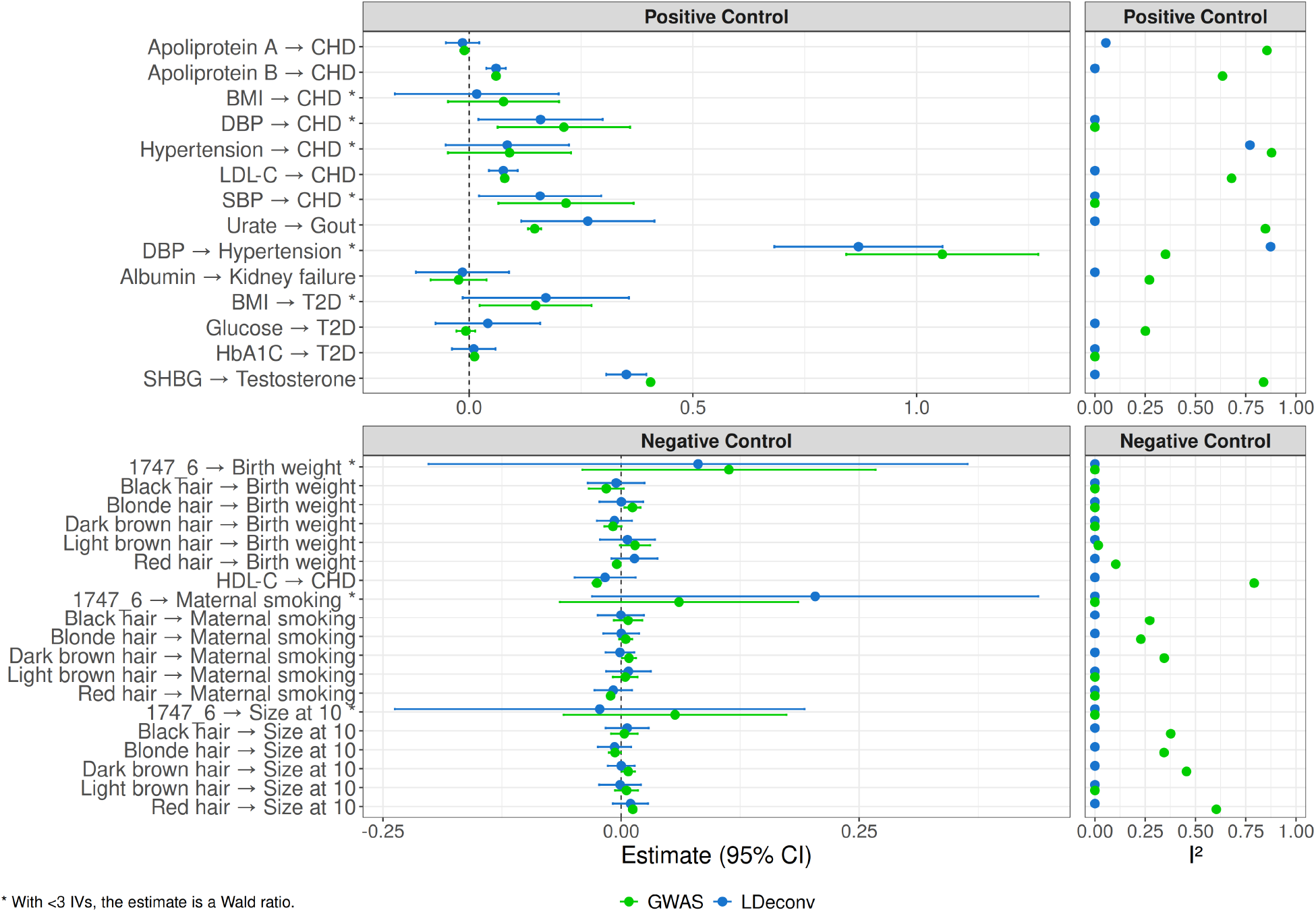
Forest plot of Mendelian randomization results for positive and negative controls. Tested exposure-outcome pairs are shown on the y-axis. The left panel shows IVW causal effect estimates with 95% confidence intervals, while the right panel shows the corresponding I^2^ values. Results obtained from GWAS are in green and those obtained from LDeconv are in blue. Positive and negative controls are displayed in separate panels. * indicates exposure-outcome pairs with <3 IVs.

For the positive controls, GWAS detected statistically significant associations in 10 out of 14 tested pairs, but with evidence of heterogeneity measured by I2 in 9 pairs, whereas LDeconv identified significant associations in half of the pairs, but heterogeneity was observed only in 3 pairs. For the negative controls, GWAS identified false positive relationships in 4 out of 10 pairs, and heterogenity in the causal estimate in 4 pairs. In contrast, LDeconv did not identify any statistically significant relationship or evidence of heterogeneity across the 10 negative controls, suggesting a lower propensity for false-positive findings.

We summarized performance with a PR-AUC derived from precision-recall curves (Supplementary Figure 5 and Methods), yielding 0.667 for GWAS and 0.806 for *LDeconv*. In addition, the *I*^2^ index was consistently lower for *LDeconv*, indicating improved concordance of Wald estimates within each exposure-outcome pair. Overall, using *LDeconv* summary statistics substantially reduced the risk of false positives in MR, at the cost of decreased recall. Unlike fine-mapping approaches, which mainly prioritize candidate causal variants, *LDeconv* produces adjusted summary statistics that can be directly used in downstream analyses such as MR.

## Discussion

We developed a LD-aware method based on truncated singular value decomposition that allows to adjust GWAS summary statistics using the LD matrix, called *LDeconv*. In simulations and real data, the adjusted summary statistics predicted causal variants more accurately than standard GWAS, albeit with some loss of recall. A key advantage of this approach is that it directly outputs deconvolved effect size estimates (*i*.*e*. LD-adjusted summary statistics), whereas fine-mapping methods typically assign posterior probabilities of causality without providing an adjusted effect size. We further showed that using LD-adjusted summary statistics improved the reliability of downstream post-GWAS analyses such as Mendelian randomization. The novelty of our approach lies in two main contributions. First, we explicitly adjust GWAS effect sizes to mitigate LD-driven inflation in marginal estimates. Second, we provide a practical strategy to approximate the inversion of the full UK Biobank LD matrix and make these resources available to the community.

However, *LDeconv* has important limitations. First, inverting the LD matrix is intrinsically difficult: the empirical LD matrix is typically ill-conditioned and often not invertible in the strict sense, requiring the use of a truncated pseudo-inverse, with the usual trade-offs and potential biases induced by regularization. In addition, matrix inversion at large scale is computationally expensive, while working with smaller local matrices may miss long-range LD. Thus, there is an unavoidable tension between mathematical and computational feasibility, and retention of biological information of the LD structure. Second, the precision of *LDeconv* depends on the ancestral alignment between the GWAS cohort and the LD reference panel. Sensitivity analyses show that *LDeconv* tolerates minor structural noise up to 5%. However, differences in LD architecture, such as between European and African cohorts, lead to a drop in performance (Supplementary Results, Supplementary Figures 8 and 9). This confirms that using ancestrally matched LD references is necessary for reliable deconvolved effect sizes. Third, the sign of marginal GWAS effects depends on the reference allele, which is partly arbitrary. Allele misspecification can flip effect directions and LD signs, and because deconvolution effectively subtracts contributions according to LD, such errors can readily propagate into false positives. For these reasons, we recommend restricting the results to variants that are significant in both GWAS and *LDeconv* when interpreting results. Finally, we do not claim that this method is competitive with the broad range of fine-mapping approaches designed specifically for causal variant discovery. Rather, our method should be viewed primarily as a tool to adjust inflated marginal effect sizes and to support downstream analyses that benefit from LD-adjusted summary statistics. We note, however, that fine-mapping methods often rely on multiple user-specified priors and decision points (*e*.*g*. LD window definition, credible set coverage, PIP thresholds), whereas LD *LDeconv* is comparatively less dependent on these tuning parameters.

## Data Availability

LDeconv code is available on GitHub at github.com/martintnr/LDeconv and full LD inverse matrices will be available on Zenodo upon publication.

## Data and code availability

*LDeconv* code is available on GitHub at github.com/martintnr/LDeconv and full LD inverse matrices will be available on Zenodo upon publication.

A benchmark of the inversion time is provided in the Supplementary Results and Supplementary Figure 10.

