## Supplementary Information for "Deriving LD-adjusted GWAS summary statistics through linkage disequilibrium deconvolution"

#### Supplementary Figures

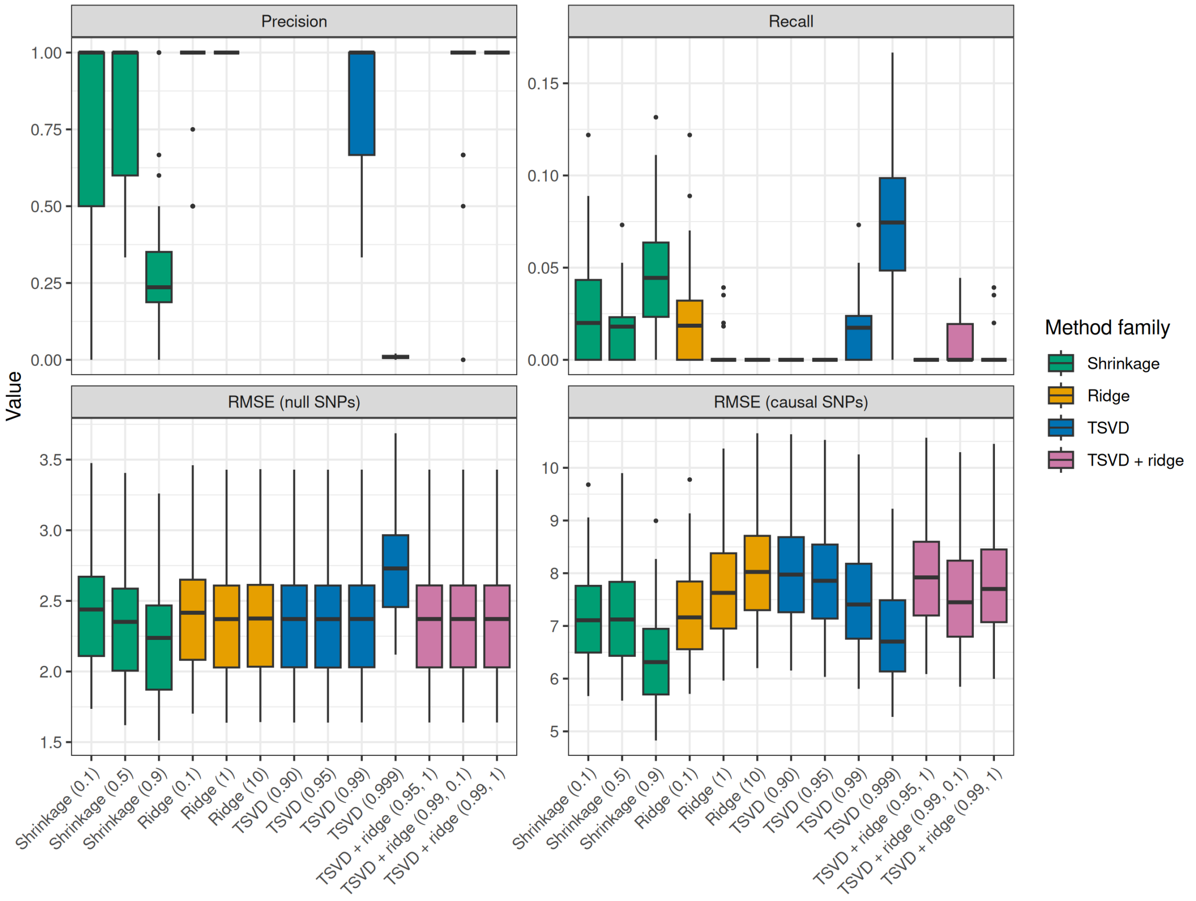


***Supplementary Figure 1.*** *Performance of LD matrix pseudo-inversion methods in simulations on chromosome 22, with low polygenicity and low heritability. The top panels show precision and recall for identifying causal variants at the genome-wide significance threshold* $p < 5 \times 10^{-8}$*. The bottom panels show the RMSE for non-causal and causal variants. The x-axis lists methods grouped and colored by inversion family: shrinkage, ridge inversion, truncated singular value decomposition (TSVD), and TSVD followed by ridge inversion. Values in parentheses indicate the shrinkage or ridge hyperparameter, the retained variance threshold for TSVD, or the retained variance threshold and ridge hyperparameter, respectively. Each boxplot summarizes 50 simulation replicates performed*

with a heritability of 0.5, polygenicity of 0.001, and GWAS sample size of 500,000. Red dots represent the mean. Precision is shown only for replicates yielding at least one significant variant.*

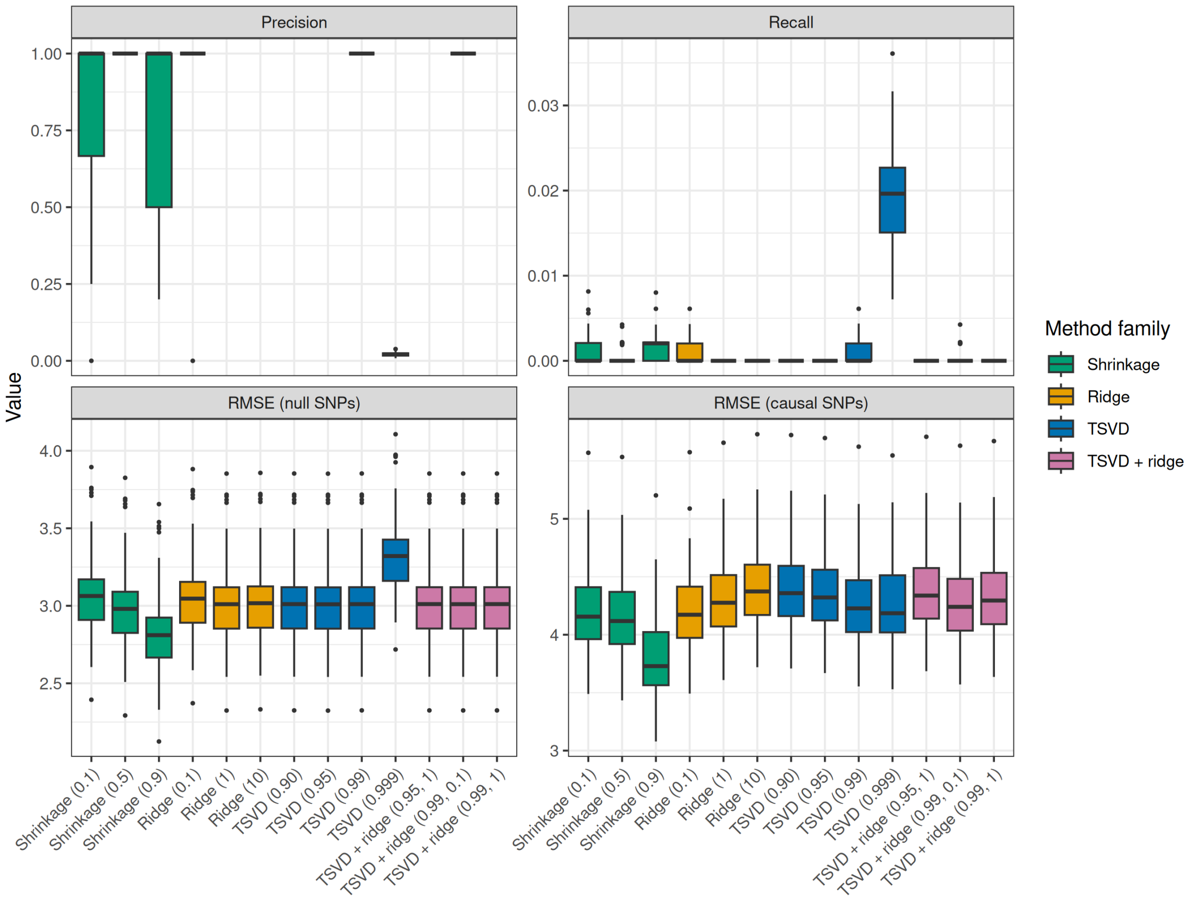


***Supplementary Figure 2.*** *Performance of LD matrix pseudo-inversion methods in simulations on chromosome 22, with high polygenicity and high heritability. The top panels show precision and recall for identifying causal variants at the genome-wide significance threshold $p < 5 \times 10^{-8}$. The bottom panels show the RMSE for non-causal and causal variants. The x-axis lists methods grouped and colored by inversion family: shrinkage, ridge inversion, truncated singular value decomposition (TSVD), and TSVD followed by ridge inversion. Values in parentheses indicate the shrinkage or ridge hyperparameter, the retained variance threshold for TSVD, or the retained variance threshold and ridge hyperparameter, respectively. Each boxplot summarizes 50 simulation replicates performed with a heritability of 0.8, polygenicity of 0.01, and GWAS sample size of 500,000. Red dots represent the mean. Precision is shown only for replicates yielding at least one significant variant.*

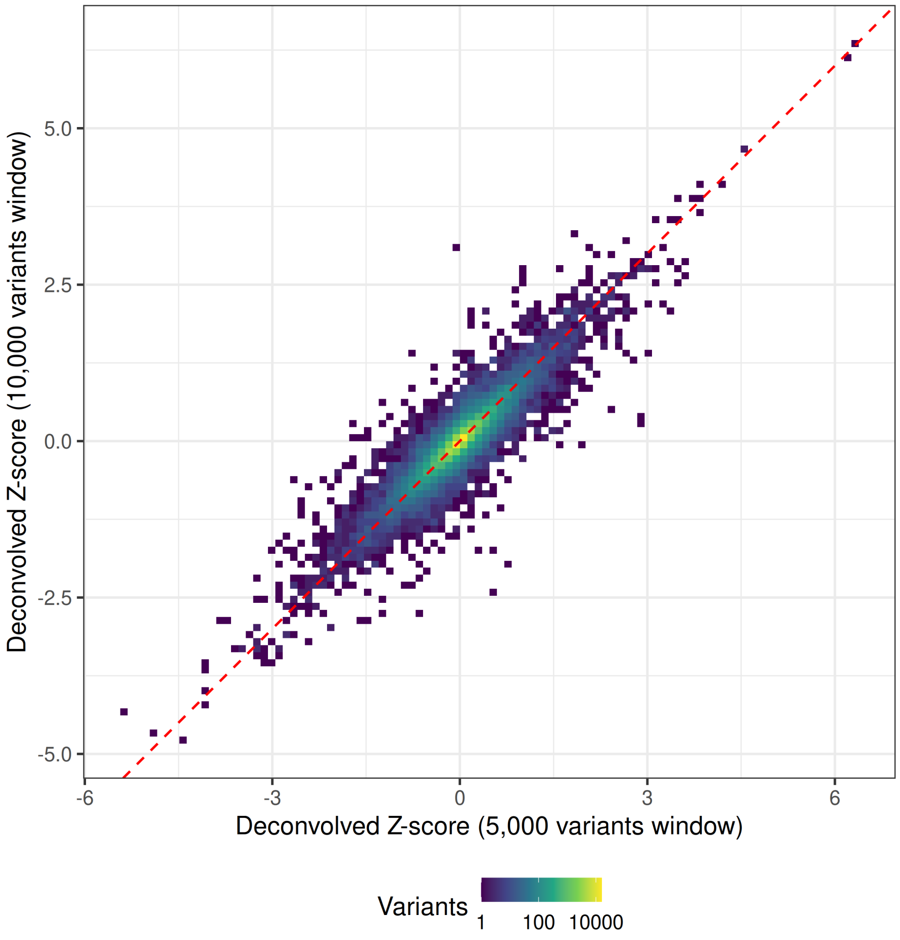


***Supplementary Figure 3.*** *Comparison of deconvolved Z-scores obtained using LDeconv windows containing fewer than 5,000 or 10,000 variants for triglyceride summary statistics on chromosome 19. The x- and y-axes show the Z-scores obtained with windows of 5,000 and 10,000 variants, respectively. Each two-dimensional bin allows to group variants with similar Z-scores, and the color indicates the number of variants within each bin on a logarithmic scale. The dashed red line represents the identity line, along which the Z-scores obtained with the two window sizes are equal.*

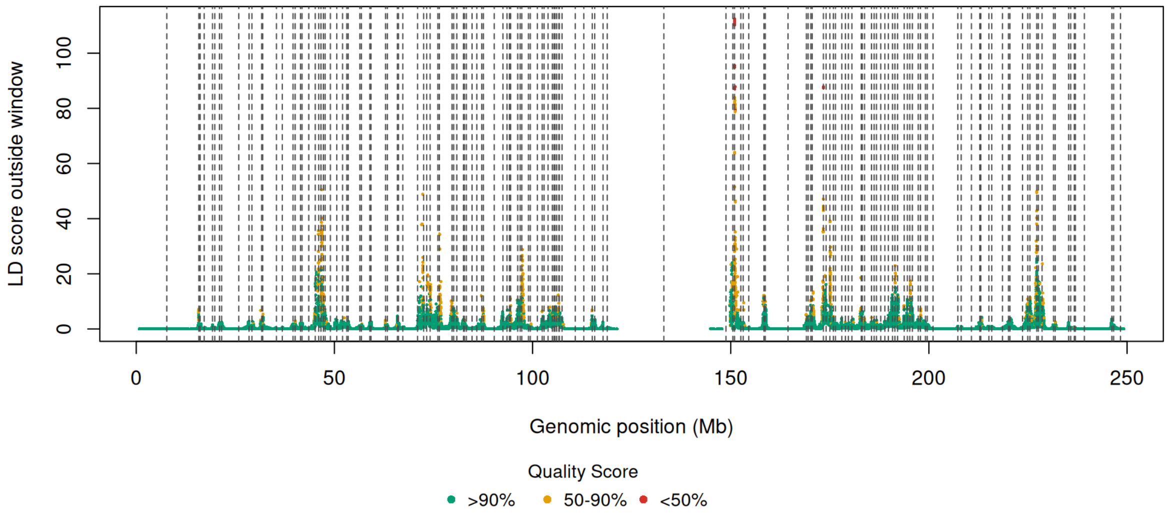


***Supplementary Figure 4.*** *Quality score for chromosome 1. Each point represents a variant, plotted by genomic position in Mb on the x-axis. Quality Score corresponds to the percentage of the total LD score captured within the window. Dashed vertical lines indicate LD block boundaries.*

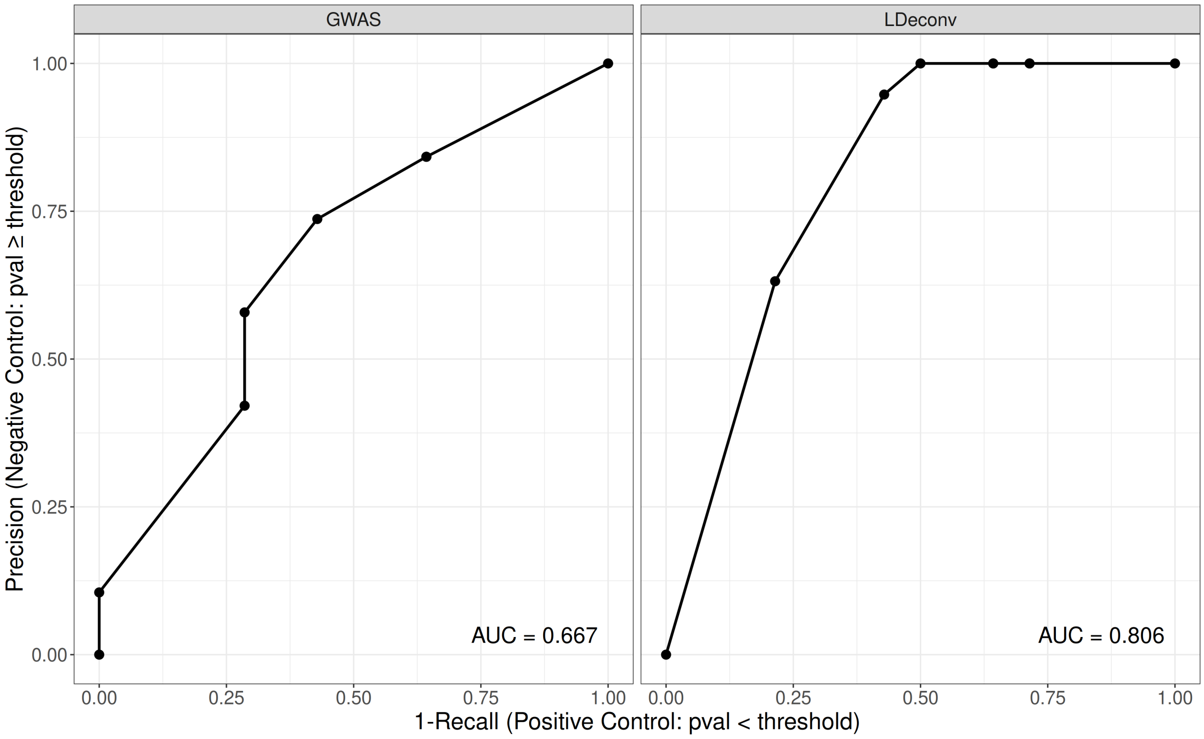


***Supplementary Figure 5.*** *Precision-(1-recall) curves for the MR positive-control and negative-control analyses across different p-value thresholds LDeconv (left) and GWAS (right). The x-axis shows 1-recall for positive controls (p-value < threshold), and the y-axis shows precision for negative controls (p-value ≥ threshold). Points denote the average precision and 1-recall for individual thresholds, and lines connect values across thresholds. The area under the curve (AUC) is reported in each panel, with higher values indicating better overall performance.*

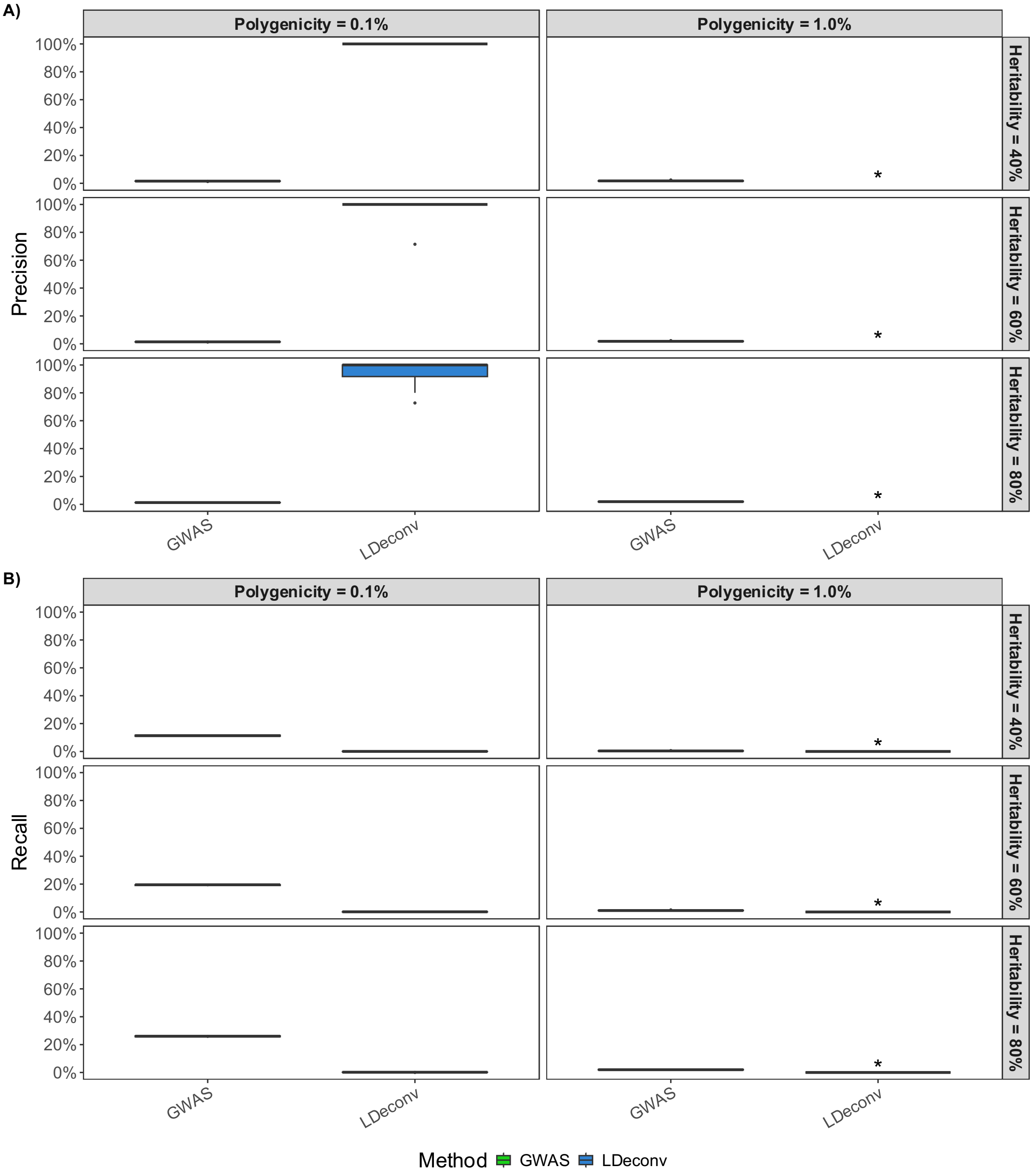


***Supplementary Figure 6.*** *A) Precision and B) Recall of predicted causal variants in simulations. The x-axis specifies method: green for the observed GWAS results and blue for LDeconv. For a given genetic variant, statistical significance was defined as $P < 5 \times 10^{-8}$ for GWAS and LDeconv. Results are shown across six simulation scenarios (panels) spanning combinations of polygenicity in columns and heritability in rows. Here, polygenicity denotes the proportion of independent variants that are truly causal to the simulated trait, and heritability is the proportion of trait variance explained by genetic effects. Each boxplot summarizes the performance over 10 independent replicates. Simulated sample size is $100{,}000$. A star above a boxplot indicates that the method did not predict any significant variants in that scenario. Consequently, precision is undefined and recall is 0.*

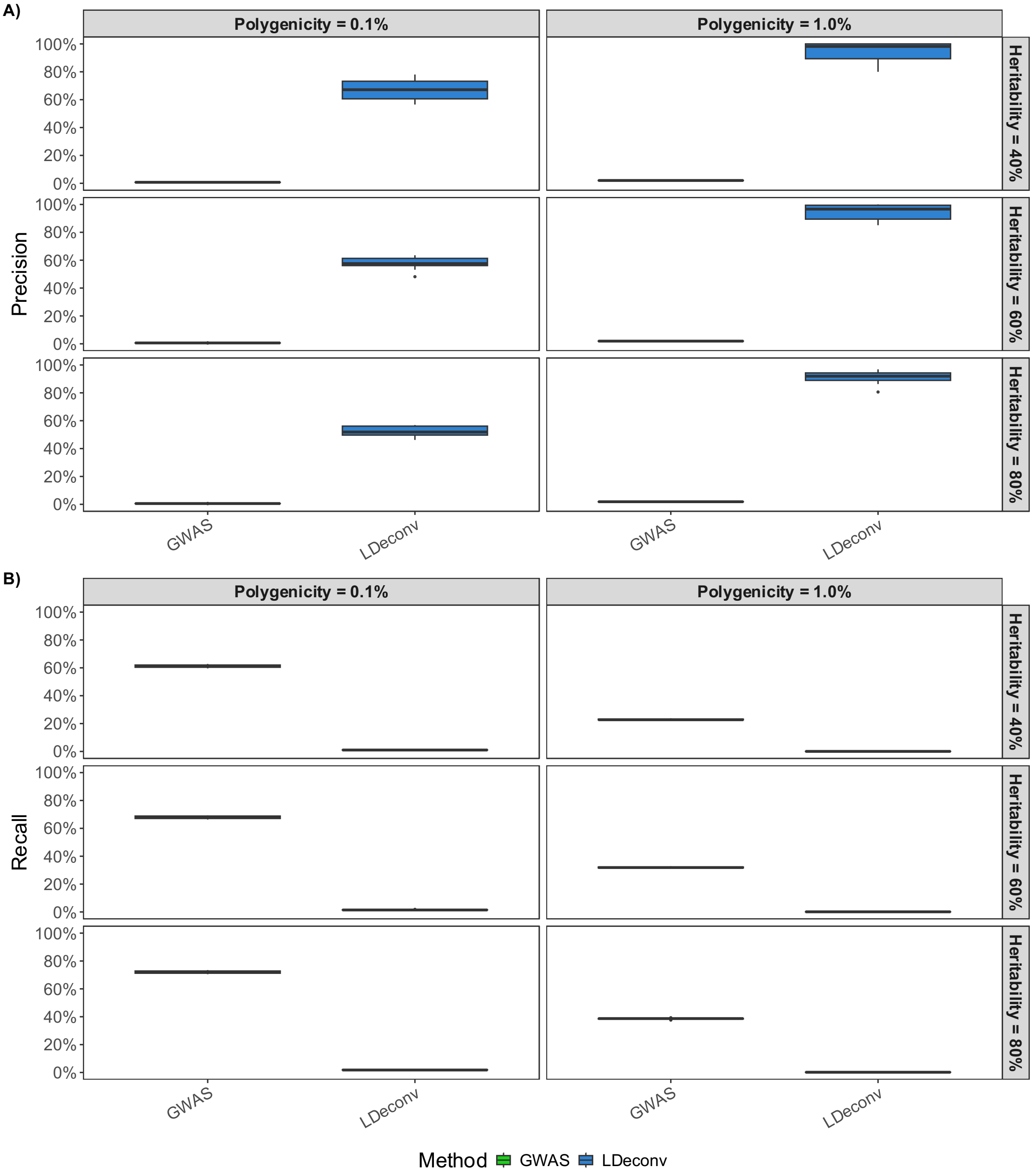


***Supplementary Figure 7.*** *A) Precision and B) Recall of predicted causal variants in simulations. The x-axis specifies the method: green for the observed GWAS results and blue for LDeconv. For a given genetic variant, statistical significance was defined as* $P < 5 \times 10^{-8}$ *for GWAS and deconvolution. Results are shown across six simulation scenarios (panels) spanning combinations of polygenicity in columns and heritability in rows. Here, polygenicity denotes the proportion of independent variants that are truly causal to the simulated trait, and heritability is the proportion of trait variance explained by genetic effects. Each boxplot summarizes the performance over 10 independent replicates. Simulated sample size is* $1{,}000{,}000$.

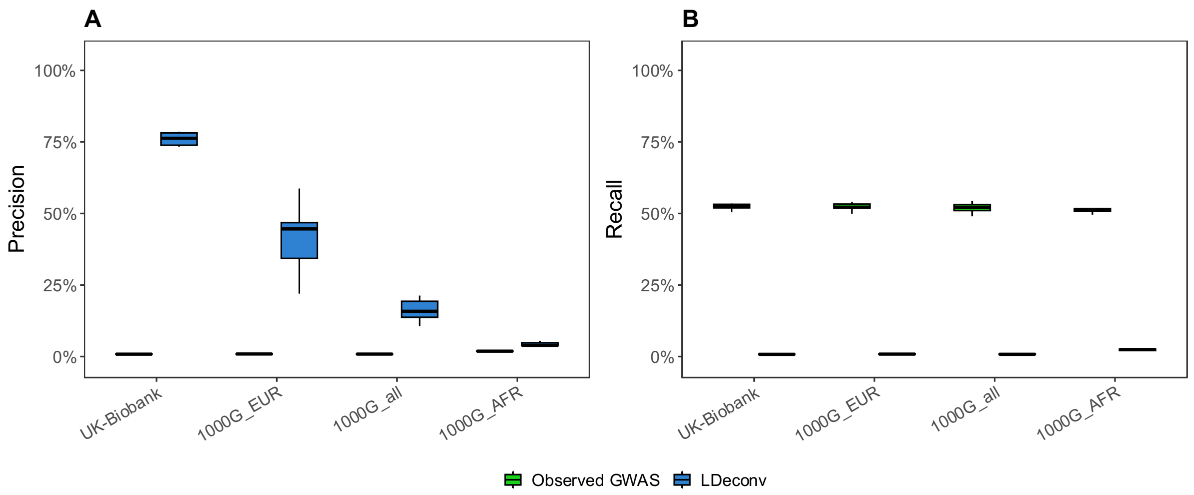


***Supplementary Figure 8.*** *Sensitivity of LDeconv precision to ancestral mismatch in LD reference. The figure displays the precision of causal variant identification across three 1000 Genomes (1000G) population subsets (EUR, ALL, AFR) using UK Biobank (UKB) inverse LD matrices for deconvolution. Precision is defined as the proportion of true causal variants among those reaching genome-wide significance* ($P < 5 \times 10^{-8}$) *post-adjustment*

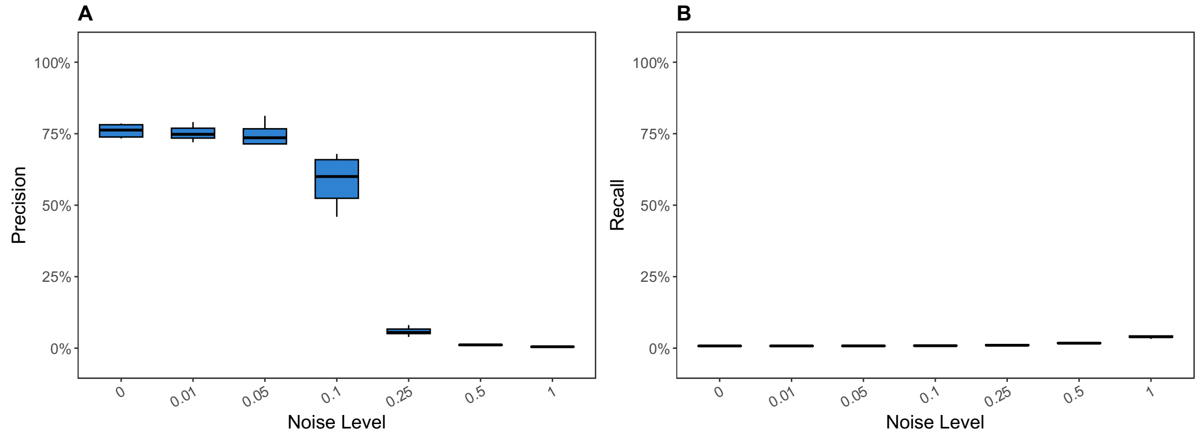


***Supplementary Figure 9.*** *Sensitivity of LDeconv precision to structural noise in the inverse LD matrix. The y-axis represents the precision across 10 independent replicates, i.e. the ratio of true causal variants to identified significant variants. The x axis represent the noise level* $1 - a$*, with* $a$ *the interpolation parameter between 0 and 1 used to build the LD matrix as* $aLD + (1 - a)LD_{rot180}$*, where* $LD_{rot180}$ *is a 180-degree rotation of the LD matrix. A value of* $a = 1$ *represents the noise-free reference LD used in LDeconv, while* $a = 0$ *represents maximal structural noise.*

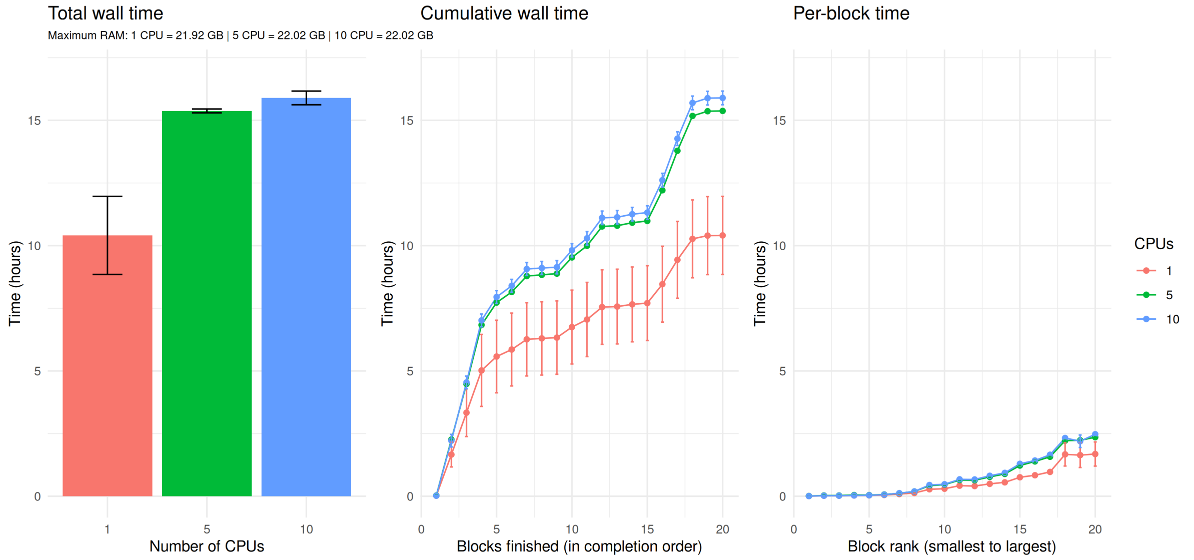


***Supplementary Figure 10.*** *Benchmark of LD matrix inversion for chromosome 21. Total wall time, cumulative wall time as LD blocks are completed, and individual per-block inversion time. Computation was performed using 1, 5, and 10 CPUs, shown in red, green, and blue, respectively. In the first panel, bars represent the mean total wall time across three independent runs, with error bars indicating standard deviation. Maximum RAM usage was 16.97 GB, 17.02 GB, and 17.00 GB for 1, 5, and 10 CPUs, respectively. In the second panel, blocks are ordered by completion, and wall time corresponds to the elapsed time at each successive block completion. In the third panel, blocks are ordered from smallest to largest according to block size, and the time required to invert each block is shown. Points represent the mean across three independent runs, with error bars indicating standard deviation.*

### Supplementary Tables

***Supplementary Table 1.*** *Deviations from Moore-Penrose properties for inverse LD matrices, summarized by chromosome.*

| Chromosome | Total number of blocks | Median error property 1 | Worse block error property 1 | Median error Frobenius | Worse block error Frobenius | Median error property 2 | Worse block error property 2 | Median error property 3 | Worse block error property 3 | Median error property 4 | Worse block error property 4 |
|---|---|---|---|---|---|---|---|---|---|---|---|
| 1 | 281 | 0.2054 | 0.5733 | 0.004127 | 0.005102 | 5.995e-15 | 3.197e-14 | 0.002365 | 0.01436 | 3.109e-15 | 2.275e-14 |
| 2 | 335 | 0.1889 | 0.4872 | 0.003756 | 0.005352 | 5.773e-15 | 2.043e-14 | 0.001917 | 0.01767 | 3.22e-15 | 2.655e-14 |
| 3 | 256 | 0.2144 | 0.7564 | 0.0039055 | 0.01072 | 5.329e-15 | 2.354e-14 | 0.0019245 | 0.01284 | 3.109e-15 | 1.932e-14 |
| 4 | 348 | 0.24095 | 0.8281 | 0.004042 | 0.005571 | 3.997e-15 | 3.952e-14 | 0.0007691 | 0.01302 | 2.702e-15 | 1.815e-14 |
| 5 | 312 | 0.20795 | 0.8726 | 0.003912 | 0.005297 | 4.663e-15 | 2.354e-14 | 0.0008028 | 0.01376 | 2.939e-15 | 1.354e-14 |
| 6 | 190 | 0.2256 | 0.6547 | 0.0039545 | 0.005073 | 5.9395e-15 | 2.709e-14 | 0.0023635 | 0.01461 | 3.0745e-15 | 7.515e-15 |
| 7 | 197 | 0.1994 | 0.6408 | 0.003889 | 0.008652 | 6.883e-15 | 2.265e-14 | 0.002627 | 0.01346 | 3.22e-15 | 2.272e-14 |
| 8 | 148 | 0.2155 | 0.5559 | 0.0038285 | 0.005728 | 7.105e-15 | 3.553e-14 | 0.002828 | 0.009961 | 3.473e-15 | 1.893e-14 |
| 9 | 122 | 0.1729 | 0.4146 | 0.0038085 | 0.004612 | 8.327e-15 | 3.642e-14 | 0.0033345 | 0.0145 | 3.5195e-15 | 1.647e-14 |
| 10 | 163 | 0.2105 | 0.7919 | 0.003904 | 0.005179 | 6.661e-15 | 2.887e-14 | 0.002361 | 0.01626 | 3.442e-15 | 1.134e-14 |
| 11 | 243 | 0.1881 | 0.9343 | 0.003858 | 0.08254 | 4.996e-15 | 2.442e-14 | 0.001044 | 0.01455 | 2.92e-15 | 1.433e-14 |
| 12 | 288 | 0.1963 | 0.6515 | 0.00413 | 0.006169 | 4.3925e-15 | 2.554e-14 | 0.0004841 | 0.01478 | 2.716e-15 | 1.222e-14 |
| 13 | 153 | 0.2147 | 0.7048 | 0.004234 | 0.005416 | 4.885e-15 | 2.931e-14 | 0.001857 | 0.01559 | 2.831e-15 | 9.159e-15 |
| 14 | 101 | 0.2011 | 0.6306 | 0.003888 | 0.004802 | 7.55e-15 | 2.132e-14 | 0.003023 | 0.01537 | 3.442e-15 | 1.497e-14 |
| 15 | 147 | 0.1548 | 0.5043 | 0.003719 | 0.006138 | 6.439e-15 | 2.398e-14 | 0.00174 | 0.01639 | 3.545e-15 | 1.287e-14 |
| 16 | 104 | 0.1468 | 0.2559 | 0.0037025 | 0.005042 | 9.437e-15 | 3.464e-14 | 0.0035265 | 0.01428 | 3.9195e-15 | 1.097e-14 |
| 17 | 90 | 0.16315 | 0.6834 | 0.003629 | 0.005375 | 7.772e-15 | 3.02e-14 | 0.0025 | 0.01551 | 3.83e-15 | 3.77e-14 |
| 18 | 90 | 0.1877 | 0.5441 | 0.0041315 | 0.005261 | 7.55e-15 | 2.709e-14 | 0.002557 | 0.0128 | 3.053e-15 | 7.861e-15 |
| 19 | 106 | 0.161 | 0.705 | 0.004218 | 0.005308 | 5.551e-15 | 3.286e-14 | 0.0015155 | 0.01448 | 2.9925e-15 | 8.865e-15 |
| 20 | 62 | 0.17075 | 0.3554 | 0.0040205 | 0.005002 | 9.881e-15 | 2.931e-14 | 0.004435 | 0.01666 | 3.525e-15 | 1.118e-14 |
| 21 | 32 | 0.19115 | 0.2849 | 0.004112 | 0.004785 | 1.2325e-14 | 2.176e-14 | 0.004375 | 0.01158 | 3.525e-15 | 7.05e-15 |
| 22 | 46 | 0.15545 | 0.2904 | 0.0039515 | 0.005041 | 8.438e-15 | 2.132e-14 | 0.004771 | 0.01345 | 3.747e-15 | 1.373e-14 |

***Supplementary Table 2.*** *Parameters of all scenarios used to simulate GWAS summary statistics.*

| **Heritability** | **Polygenicity** | **Sample size** |
|---|---|---:|
| 0.4 | 0.001 | 500,000 |
| 0.4 | 0.001 | 100,000 |
| 0.4 | 0.001 | 1,000,000 |
| 0.4 | 0.01 | 500,000 |
| 0.4 | 0.01 | 100,000 |
| 0.4 | 0.01 | 1,000,000 |
| 0.6 | 0.001 | 500,000 |
| 0.6 | 0.001 | 100,000 |
| 0.6 | 0.001 | 1,000,000 |
| 0.6 | 0.01 | 500,000 |
| 0.6 | 0.01 | 100,000 |
| 0.6 | 0.01 | 1,000,000 |
| 0.8 | 0.001 | 500,000 |
| 0.8 | 0.001 | 100,000 |
| 0.8 | 0.001 | 1,000,000 |
| 0.8 | 0.01 | 500,000 |
| 0.8 | 0.01 | 100,000 |
| 0.8 | 0.01 | 1,000,000 |

***Supplementary Table 3.*** *Description of the traits included from the UK Biobank. The first column specifies the trait category, the following four columns correspond to the Trait code in UK Biobank, the description of the trait in UK Biobank, the trait name we used throughout the paper, and the sample size. The final column specifies the analysis in which the traits were used.*

| Category | Trait code in UKBiobank | Description of the trait in UK Biobank | Trait name | Sample size | Analysis |
|---|---|---|---|---|---|
| Anthropometric trait | 21001_irnt | Body mass index (BMI) | BMI | 359,983 | General results |
| Anthropometric trait | 23099_irnt | Body fat percentage | Body fat percentage | 354,628 | General results |
| Anthropometric trait | 23106_irnt | Impedance of whole body | Body impedance | 354,795 | General results |
| Anthropometric trait | 3148_irnt | Heel bone mineral density (BMD) | BMD | 206,496 | General results |
| Anthropometric trait | 48_irnt | Waist circumference | Waist circumference | 360,564 | General results |
| Anthropometric trait | 50_irnt | Standing height | Standing height | 360,388 | General results |
| Biomarker | 30600_irnt | Albumin (g/L) | Albumin | 315,268 | General results |
| Biomarker | 30620_irnt | Alanine aminotransferase (U/L) | Alanine aminotransferase | 344,136 | General results |
| Biomarker | 30650_irnt | Aspartate aminotransferase (U/L) | Aspartate aminotransferase | 342,990 | General results |
| Biomarker | 30680_irnt | Calcium (mmol/L) | Calcium | 315,153 | General results |
| Biomarker | 30700_irnt | Creatinine (umol/L) | Creatinine | 344,104 | General results |
| Biomarker | 30710_irnt | C-reactive protein (mg/L) | CRP | 343,524 | General results |
| Biomarker | 30730_irnt | Gamma glutamyltransferase (U/L) | Gamma glutamyltransferase | 344,104 | General results |
| Biomarker | 30740_irnt | Glucose (mmol/L) | Glucose | 314,916 | General results |
| Biomarker | 30750_irnt | Glycated haemoglobin (mmol/mol) | HbA1C | 344,182 | General results |
| Biomarker | 30770_irnt | IGF-1 (nmol/L) | IGF-1 | 342,439 | General results |
| Biomarker | 30810_irnt | Phosphate (mmol/L) | Phosphate | 314,658 | General results |
| Biomarker | 30850_irnt | Testosterone (nmol/L) | Testosterone | 312,102 | General results |
| Biomarker | 30860_irnt | Total protein (g/L) | Total protein | 314,921 | General results |
| Biomarker | 30880_irnt | Urate (umol/L) | Urate | 343,836 | General results |
| Biomarker | 30890_irnt | Vitamin D (nmol/L) | Vitamin D | 329,247 | General results |
| Blood trait | 30150 | Eosinophill count | Eosinophil count | 349,856 | General results |
| Blood trait | 30160 | Basophill count | Basophil count | 349,856 | General results |
| Blood trait | 30000_irnt | White blood cell (leukocyte) count | White blood cell count | 350,470 | General results |
| Blood trait | 30010_irnt | Red blood cell (erythrocyte) count | Red blood cell count | 350,475 | General results |
| Blood trait | 30080_irnt | Platelet count | Platelet count | 350,474 | General results |
| Blood trait | 30120_irnt | Lymphocyte count | Lymphocyte count | 349,856 | General results |
| Blood trait | 30130_irnt | Monocyte count | Monocyte count | 349,856 | General results |
| Blood trait | 30140_irnt | Neutrophill count | Neutrophil count | 349,856 | General results |
| Blood trait | 30180_irnt | Lymphocyte percentage | Lymphocyte percentage | 349,861 | General results |
| Blood trait | 30250_irnt | Reticulocyte count | Reticulocyte count | 344,729 | General results |
| Cancer trait | C_PANCREAS | Malignant neoplasm of pancreas | Pancreas cancer | 361,194 | General results |
| Cancer trait | C3_SKIN | Malignant neoplasm of skin | Skin cancer | 361,194 | General results |
| Cancer trait | C50 | Diagnoses - main ICD10: C50 Malignant neoplasm of breast | Breast cancer | 361,194 | General results |
| Cancer trait | II_NEOPLASM | Neoplasms | Cancer | 361,194 | General results |
| Education | 20023_irnt | Mean time to correctly identify matches | Match identification exercise | 358,695 | General results |
| Education | 6138_1 | Qualifications: College or University degree | University degree | 357,549 | General results |
| Health | 2178 | Overall health rating | Overall health rating | 359,681 | General results |
| Inflammatory trait | K51 | Diagnoses - main ICD10: K51 Ulcerative colitis | UC | 361,194 | General results |
| Lifestyle trait | 884 | Number of days/week of moderate physical activity 10+ minutes | Physical activity | 343,943 | General results |
| Lifestyle trait | 1200 | Sleeplessness / insomnia | Insomnia | 360,738 | General results |
| Lifestyle trait | 1458 | Cereal intake | Cereal intake | 345,019 | General results |
| Lifestyle trait | 1558 | Alcohol intake frequency. | Alcohol freq | 360,726 | General results |

| Category | Trait code in UKBiobank | Description of the trait in UK Biobank | Trait name | Sample size | Analysis |
|---|---|---|---|---|---|
| Lifestyle trait | 1468_4 | Cereal type: Muesli | Muesli intake | 299,898 | General results |
| Lifestyle trait | 20116_0 | Smoking status: Never | Never smoker | 359,706 | General results |
| Lipid | 30630_irnt | Apoliprotein A (g/L) | Apoliprotein A | 313,387 | General results |
| Lipid | 30640_irnt | Apoliprotein B (g/L) | Apoliprotein B | 342,590 | General results |
| Lipid | 30690_irnt | Cholesterol (mmol/L) | Cholesterol | 344,278 | General results |
| Lipid | 30760_irnt | HDL cholesterol (mmol/L) | HDL-C | 315,133 | General results |
| Lipid | 30780_irnt | LDL direct (mmol/L) | LDL-C | 343,621 | General results |
| Lipid | 30870_irnt | Triglycerides (mmol/L) | Triglycerides | 343,992 | General results |
| Metabolic trait | 20002_1065 | Non-cancer illness code, self-reported: hypertension | Hypertension | 361,141 | General results |
| Metabolic trait | 20002_1223 | Non-cancer illness code, self-reported: type 2 diabetes | T2D | 361,141 | General results |
| Metabolic trait | 20002_1466 | Non-cancer illness code, self-reported: gout | Gout | 361,141 | General results |
| Metabolic trait | 23105_irnt | Basal metabolic rate | Basal metabolic rate | 354,825 | General results |
| Metabolic trait | 4079_irnt | Diastolic blood pressure, automated reading | DBP | 340,162 | General results |
| Metabolic trait | I25 | Diagnoses - main ICD10: I25 Chronic ischaemic heart disease | CHD | 361,194 | General results |
| Psychiatric trait | 20127_irnt | Neuroticism score | Neuroticism score | 293,006 | General results |
| Psychiatric trait | V_MENTAL_BEHAV | Mental and behavioural disorders | Mental disorders | 361,194 | General results |
| Respiratory trait | 20002_1111 | Non-cancer illness code, self-reported: asthma | Asthma | 361,141 | General results |
| Respiratory trait | 3062_irnt | Forced vital capacity (FVC) | Forced vital capacity | 329,404 | General results |
| Health | 1757 | Facial ageing | Facial ageing | 330,409 | General results |
| Lifestyle trait | 1528 | Water intake | Water intake | 333,363 | General results |
| Biomarker | 30670_irnt | Urea (mmol/L) | Urea | 344,052 | General results |
| Lifestyle trait | 1488_irnt | Tea intake | Tea intake | 349,376 | General results |
| Blood trait | 30070_irnt | Red blood cell (erythrocyte) distribution width | Red blood cell DW | 350,473 | General results |
| Lifestyle trait | 6155_100 | Vitamin and mineral supplements: None of the above | No vitamins | 359,245 | General results |
| Biomarker | 30830_irnt | SHBG (nmol/L) | SHBG | 312,215 | General results |
| Biomarker | 30610_irnt | Alkaline phosphatase (U/L) | Alkaline phosphatase | 344,292 | General results |
| Biomarker | 30720_irnt | Cystatin C (mg/L) | Cystatin C | 344,264 | General results |
| Metabolic trait | 4080_irnt | Systolic blood pressure, automated reading | SBP | 340,159 | Mendelian randomization |
| Metabolic trait | 20002_1192 | Non-cancer illness code, self-reported: renal/kidney failure | Kidney failure | 361,141 | Mendelian randomization |
| Hair color | 1747_1 | Hair colour (natural, before greying): Blonde | Blonde hair | 360,270 | Mendelian randomization |
| Hair color | 1747_2 | Hair colour (natural, before greying): Red | Red hair | 360,270 | Mendelian randomization |
| Hair color | 1747_3 | Hair colour (natural, before greying): Light brown | Light brown hair | 360,270 | Mendelian randomization |
| Hair color | 1747_4 | Hair colour (natural, before greying): Dark brown | Dark brown hair | 360,270 | Mendelian randomization |
| Hair color | 1747_5 | Hair colour (natural, before greying): Black | Black hair | 360,270 | Mendelian randomization |
| Childhood trait | 1687 | Comparative body size at age 10 | Size at 10 | 354,996 | Mendelian randomization |
| Childhood trait | 1787 | Maternal smoking around birth | Maternal smoking | 309,942 | Mendelian randomization |
| Childhood trait | 20022_irnt | Birth weight | Birth weight | 205,475 | Mendelian randomization |

***Supplementary Table 4.*** *Full results of the positive and negative control Mendelian randomization analyses. The table reports the results of IVW MR analyses for the positive and negative controls using GWAS and LDeconv estimates. The columns show, in order: the IVW causal effect estimate, the standard error of the IVW estimate, the significance of the IVW test, Cochran's Q statistic for heterogeneity across instrumental variables, the degrees of freedom for the Q statistic, the proportion of between-instrument heterogeneity I2, the analysis framework used, the exposure trait code, the outcome trait code, the number of instrumental variables included in the analysis, and whether the analysis corresponds to a positive or negative control.*

| IVW estimate | standard error | p-value | Q-statistic | Q degrees of freedom | I2 | Condition | Exposure | Outcome | Number of IVs | Control |
|---|---|---|---|---|---|---|---|---|---|---|
| 0.0789874708883902 | 0.00404705044434209 | 7.82764589500584e-85 | 84.0762638987401 | 27 | 0.678862990004904 | GWAS | 30780_irnt | I25 | 28 | Positive Control |
| 0.0732311948431293 | 0.0161164928337112 | 5.52315967533645e-06 | 3.34785057874609 | 27 | 0 | LDeconv | 30780_irnt | I25 | 28 | Positive Control |
| 0.0598922604494435 | 0.00277076985828921 | 1.27718894770602e-103 | 111.74372996355 | 44 | 0.60624188923752 | GWAS | 30640_irnt | I25 | 45 | Positive Control |
| 0.0568391386203659 | 0.0109879905122676 | 2.3055998880811e-07 | 6.27591976915596 | 44 | 0 | LDeconv | 30640_irnt | I25 | 45 | Positive Control |
| 0.184618495386812 | 0.0546500721151281 | 0.000729638309398693 | 71.0717588101906 | 4 | 0.943718854479419 | GWAS | 4080_irnt | I25 | 5 | Positive Control |
| 0.131794624687185 | 0.0575072836162831 | 0.0219177568042538 | 20.3591533848175 | 4 | 0.803528176030004 | LDeconv | 4080_irnt | I25 | 5 | Positive Control |
| -0.0149843942956227 | 0.00422824120123114 | 0.000394279401981068 | 261.584306854409 | 42 | 0.839439909430897 | GWAS | 30630_irnt | I25 | 43 | Positive Control |
| -0.0138126547258953 | 0.0182101682454415 | 0.448143756741302 | 36.7682027781489 | 42 | 0 | LDeconv | 30630_irnt | I25 | 43 | Positive Control |
| 0.0124315935660657 | 0.00405866781280112 | 0.00219149175296092 | 34.3815039560675 | 30 | 0.127437821267684 | GWAS | 30750_irnt | 20002_1223 | 31 | Positive Control |
| 0.0206754262761629 | 0.022710168057707 | 0.36260952532284 | 11.3508307208694 | 30 | 0 | LDeconv | 30750_irnt | 20002_1223 | 31 | Positive Control |
| 0.402109781851062 | 0.00319730451134952 | 0 | 256.90189277776 | 42 | 0.836513466110064 | GWAS | 30830_irnt | 30850_irnt | 43 | Positive Control |
| 0.333394968854623 | 0.0215988291815495 | 9.40433481745892e-54 | 51.4392835482093 | 42 | 0.183503402401839 | LDeconv | 30830_irnt | 30850_irnt | 43 | Positive Control |
| 0.154823539876736 | 0.0068517555089717 | 4.72463274187098e-113 | 54.9418688002375 | 8 | 0.854391556481504 | GWAS | 30880_irnt | 20002_1466 | 9 | Positive Control |
| 0.155284894851012 | 0.0494170054051466 | 0.0016760488519606 | 5.38246140860496 | 8 | 0 | LDeconv | 30880_irnt | 20002_1466 | 9 | Positive Control |
| -0.019778099523356 | 0.0293259860473611 | 0.500042856946074 | 7.90790377806231 | 7 | 0.114809664298265 | GWAS | 30600_irnt | 20002_1192 | 8 | Positive Control |
| -0.0568731705756563 | 0.0468192533732836 | 0.224465672784509 | 4.80411685347442 | 7 | 0 | LDeconv | 30600_irnt | 20002_1192 | 8 | Positive Control |
| 0.0116080050852338 | 0.00425981735914654 | 0.0064301359898345 | 27.7762808021718 | 39 | 0 | GWAS | 1747_1 | 20022_irnt | 40 | Negative Control |
| 0.00193311409363164 | 0.0116933463133455 | 0.86869413146675 | 16.6162864973061 | 39 | 0 | LDeconv | 1747_1 | 20022_irnt | 40 | Negative Control |
| -0.00374328350656274 | 0.00139128501970255 | 0.00713402460962969 | 146.398944471495 | 131 | 0.105184805307756 | GWAS | 1747_2 | 20022_irnt | 132 | Negative Control |
| 0.0114192614820947 | 0.011650282943376 | 0.327002055816914 | 52.3909365692961 | 131 | 0 | LDeconv | 1747_2 | 20022_irnt | 132 | Negative Control |
| 0.0152573247074836 | 0.0080931096611548 | 0.0593995995542572 | 12.9150195062151 | 13 | 0 | GWAS | 1747_3 | 20022_irnt | 14 | Negative Control |
| 0.00926161485123711 | 0.0147804608459681 | 0.530913568454223 | 7.61527224761916 | 13 | 0 | LDeconv | 1747_3 | 20022_irnt | 14 | Negative Control |
| -0.00793391152574727 | 0.00407914027676236 | 0.0517754596160891 | 19.7618322374191 | 35 | 0 | GWAS | 1747_4 | 20022_irnt | 36 | Negative Control |
| -0.00935781849706516 | 0.00945068306174628 | 0.322089189832428 | 18.0021912078396 | 35 | 0 | LDeconv | 1747_4 | 20022_irnt | 36 | Negative Control |
| -0.0147705401286046 | 0.00924804459518388 | 0.1102316556325 | 13.0461754456158 | 19 | 0 | GWAS | 1747_5 | 20022_irnt | 20 | Negative Control |
| -0.00624922082961423 | 0.0151919690348027 | 0.680815700618268 | 13.1866857039483 | 19 | 0 | LDeconv | 1747_5 | 20022_irnt | 20 | Negative Control |

| IVW estimate | standard error | p-value | Q-statistic | Q degrees of freedom | I2 | Condition | Exposure | Outcome | Number of IVs | Control |
|---|---|---|---|---|---|---|---|---|---|---|
| 0.00525402117107568 | 0.00347013479567663 | 0.130008511778079 | 50.6724638486817 | 39 | 0.230351219619754 | GWAS | 1747_1 | 1787 | 40 | Negative Control |
| 0.00321351416235271 | 0.0095303961236597 | 0.735976847359397 | 26.0925867637473 | 39 | 0 | LDeconv | 1747_1 | 1787 | 40 | Negative Control |
| -0.0112868365208096 | 0.00113089843903164 | 1.85687215104576e-23 | 137.101129344986 | 131 | 0.044500941561422 | GWAS | 1747_2 | 1787 | 132 | Negative Control |
| -0.0158730492851381 | 0.00948234993374938 | 0.0941389834412252 | 66.609937631113 | 131 | 0 | LDeconv | 1747_2 | 1787 | 132 | Negative Control |
| 0.00407955078937242 | 0.00659287021004058 | 0.536059844304443 | 6.86922216554522 | 13 | 0 | GWAS | 1747_3 | 1787 | 14 | Negative Control |
| 0.0107967725791486 | 0.0120470756270946 | 0.370137826410099 | 13.0527945818341 | 13 | 0.00404469567824141 | LDeconv | 1747_3 | 1787 | 14 | Negative Control |
| 0.00611368497122262 | 0.00332322270333559 | 0.0658144080659993 | 51.5902748700398 | 35 | 0.321577563054899 | GWAS | 1747_4 | 1787 | 36 | Negative Control |
| -0.00340588432178189 | 0.00770196193649126 | 0.658337275423979 | 31.3017330739588 | 35 | 0 | LDeconv | 1747_4 | 1787 | 36 | Negative Control |
| 0.00785995300515348 | 0.00753566254262293 | 0.296932499371334 | 24.9029755054427 | 19 | 0.237038963643222 | GWAS | 1747_5 | 1787 | 20 | Negative Control |
| -0.00142687788678604 | 0.0123831915181927 | 0.90826520612947 | 21.7776758086063 | 19 | 0.127546935357933 | LDeconv | 1747_5 | 1787 | 20 | Negative Control |
| -0.00608905087464838 | 0.00324263902512237 | 0.0604075463764661 | 56.1716774567239 | 39 | 0.30569992270488 | GWAS | 1747_1 | 1687 | 40 | Negative Control |
| -0.00873492415907668 | 0.00889933492011415 | 0.326333665678844 | 11.288318399606 | 39 | 0 | LDeconv | 1747_1 | 1687 | 40 | Negative Control |
| 0.0123231294681403 | 0.00105731460900929 | 2.16073841530086e-31 | 281.242520126455 | 131 | 0.534209834483426 | GWAS | 1747_2 | 1687 | 132 | Negative Control |
| 0.0122454960394963 | 0.00885821263505765 | 0.166852280254061 | 53.5136991477992 | 131 | 0 | LDeconv | 1747_2 | 1687 | 132 | Negative Control |
| 0.00573372926852015 | 0.00615900216816181 | 0.351878907993886 | 10.8428762500184 | 13 | 0 | GWAS | 1747_3 | 1687 | 14 | Negative Control |
| -0.00142503428605265 | 0.0112482635901867 | 0.899186339897001 | 5.57659491782334 | 13 | 0 | LDeconv | 1747_3 | 1687 | 14 | Negative Control |
| 0.00329462951477659 | 0.0031052115531124 | 0.288689892840258 | 74.2333939500526 | 35 | 0.528514080555855 | GWAS | 1747_4 | 1687 | 36 | Negative Control |
| 0.00308005659646641 | 0.00719381911400928 | 0.668539594375018 | 23.2033465134458 | 35 | 0 | LDeconv | 1747_4 | 1687 | 36 | Negative Control |
| 0.00521876298663899 | 0.00704061719750525 | 0.458550009709597 | 25.0977155492633 | 19 | 0.242958987135476 | GWAS | 1747_5 | 1687 | 20 | Negative Control |
| 0.00635537424469978 | 0.0115600590645297 | 0.582477103737401 | 8.64434648132889 | 19 | 0 | LDeconv | 1747_5 | 1687 | 20 | Negative Control |
| -0.0250651360600938 | 0.00234530682736854 | 1.16645222346487e-26 | 263.593756421628 | 55 | 0.791345588959909 | GWAS | 30760_irnt | I25 | 56 | Negative Control |
| -0.0166752371923143 | 0.0159469297192038 | 0.295713094086663 | 40.3839042756152 | 55 | 0 | LDeconv | 30760_irnt | I25 | 56 | Negative Control |

**Supplementary Table 5.** *Full data used to create Main Figure 4.*

| Trait name | Number of GWAS significant variants | Number of GWAS significant variants after clumping | Number of LDeconv significant variants |
|---|---|---|---|
| BMI | 33,141 | 465 | 5 |
| Body fat percentage | 25,910 | 399 | 2 |
| Body impedance | 45,114 | 550 | 6 |
| BMD | 31,281 | 378 | 10 |
| Waist circumference | 20,423 | 341 | 4 |
| Standing height | 120,108 | 1,293 | 47 |
| Albumin | 16,321 | 241 | 8 |
| Alanine aminotransferase | 17,291 | 243 | 11 |
| Aspartate aminotransferase | 26,518 | 295 | 19 |
| Calcium | 16,989 | 220 | 4 |
| Creatinine | 37,509 | 447 | 2 |
| CRP | 20,198 | 254 | 18 |
| Gamma glutamyltransferase | 29,090 | 361 | 34 |
| Glucose | 5,558 | 109 | 5 |
| HbA1C | 40,088 | 427 | 32 |
| IGF-1 | 51,283 | 493 | 11 |
| Phosphate | 12,863 | 174 | 5 |
| Testosterone | 6,877 | 104 | 1 |
| Total protein | 20,806 | 320 | 11 |
| Urate | 25,811 | 296 | 6 |
| Vitamin D | 6,289 | 76 | 5 |
| Eosinophil count | 33,523 | 376 | 12 |
| Basophil count | 1,651 | 32 | 4 |
| White blood cell count | 32,490 | 446 | 9 |
| Red blood cell count | 39,746 | 502 | 11 |
| Platelet count | 55,851 | 643 | 31 |
| Lymphocyte count | 35,822 | 428 | 11 |
| Monocyte count | 35,048 | 473 | 42 |
| Neutrophil count | 29,126 | 366 | 7 |
| Lymphocyte percentage | 28,735 | 362 | 10 |
| Reticulocyte count | 35,232 | 366 | 16 |
| Pancreas cancer | 0 | 0 | 2 |
| Skin cancer | 2,803 | 41 | 4 |
| Breast cancer | 665 | 14 | 2 |
| Cancer | 383 | 10 | 1 |
| Match identification exercise | 2,590 | 54 | 0 |
| University degree | 15,281 | 226 | 3 |
| Overall health rating | 3,517 | 60 | 2 |
| UC | 39 | 3 | 3 |
| Physical activity | 299 | 5 | 1 |
| Insomnia | 1,218 | 28 | 0 |
| Cereal intake | 936 | 20 | 1 |
| Alcohol freq | 5,213 | 51 | 1 |
| Muesli intake | 582 | 10 | 1 |
| Never smoker | 5,146 | 75 | 4 |
| Apoliprotein A | 24,620 | 294 | 43 |
| Apoliprotein B | 16,638 | 201 | 51 |
| Cholesterol | 17,823 | 215 | 26 |
| HDL-C | 27,951 | 341 | 61 |
| LDL-C | 14,399 | 188 | 37 |
| Triglycerides | 26,440 | 316 | 19 |
| Hypertension | 15,424 | 209 | 2 |

| Trait name | Number of GWAS significant variants | Number of GWAS significant variants after clumping | Number of LDeconv significant variants |
|---|---|---|---|
| T2D | 68 | 1 | 1 |
| Gout | 2,567 | 20 | 1 |
| Basal metabolic rate | 49,090 | 680 | 8 |
| DBP | 15,316 | 219 | 3 |
| CHD | 1,413 | 23 | 2 |
| Neuroticism score | 6,993 | 61 | 4 |
| Mental disorders | 0 | 0 | 1 |
| Asthma | 6,229 | 86 | 2 |
| Forced vital capacity | 31,206 | 386 | 12 |
| Facial ageing | 2,280 | 48 | 4 |
| Water intake | 1,042 | 22 | 1 |
| Urea | 13,899 | 186 | 3 |
| Tea intake | 2,460 | 22 | 3 |
| Red blood cell DW | 45,476 | 435 | 15 |
| No vitamins | 58 | 5 | 1 |
| SHBG | 28,339 | 342 | 50 |
| Alkaline phosphatase | 40,670 | 433 | 40 |
| Cystatin C | 34,577 | 388 | 12 |

#### Supplementary Results

##### Impact of window size on *LDeconv* results in real data

Assuming that LD between chromosomes is negligible, each chromosome can be analyzed independently. However, chromosome-wide LD matrices remain too large to invert directly. As described in the Methods section, we therefore partitioned each chromosome into adjacency-constrained LD blocks containing fewer than $10{,}000$ variants and performed deconvolution within overlapping windows. Because this block-size constraint affects both the dimensions and LD content of the matrices subjected to pseudo-inversion, we assessed the sensitivity of the results by repeating the analysis with a maximum block size of $5{,}000$ variants. We applied both settings to triglyceride summary statistics on chromosome 19. First, the classification of variants as significant or non-significant at $p < 5 \times 10^{-8}$ was identical for settings. Second, the deconvolved Z-scores showed high Pearson correlation, exceeding $0.91$. Third, as shown in Supplementary Figure 3, the Z-scores obtained using both settings were strongly correlated. We therefore conclude that *LDeconv* is reasonably robust to the choice of window size and that the small differences arising from the numerical challenges associated with LD matrix pseudo-inversion are acceptable.

##### Impact of outside-of-window LD on *LDeconv*

Because deconvolution is performed within finite windows, the LD with variants outside the deconvolution window is omitted to compute the pseudo-inverse. To assess the impact of this omission, we defined a quality score for each variant as the proportion of its chromosome-wide LD score captured within the window used for deconvolution. Defining LD score$_i = \sum_j r_{ij}^2$, where $r_{ij}$ is the genotype correlation between variants $i$ and $j$, the quality score can be written:

$$\text{Quality score} = \frac{\text{LD score of the variant within its deconvolution window}}{\text{Chromosome-wide LD score of the variant}}$$

A score close to one indicates that most of the variant's LD is captured within its window, whereas a score close to zero indicates that most is omitted and that the corresponding deconvolved Z-score should be interpreted with caution. Supplementary Figure 4 illustrates the quality scores of chromosome 1 variants. Genome-wide quality scores are included in the index available online.

##### Pseudo-inverse properties

We verified that inverse LD matrices satisfy the four defining properties of the Moore-Penrose pseudo inverse $\mathbf{LD}^+$, which are:

1. $\mathbf{LD}\,\mathbf{LD}^+\,\mathbf{LD} = \mathbf{LD}$
2. $\mathbf{LD}^+\,\mathbf{LD}\,\mathbf{LD}^+ = \mathbf{LD}^+$
3. $(\mathbf{LD}\,\mathbf{LD}^+)^\top = \mathbf{LD}\,\mathbf{LD}^+$
4. $(\mathbf{LD}^+\,\mathbf{LD})^\top = \mathbf{LD}^+\,\mathbf{LD}$

For each inverted LD block, we computed the discrepancy between the left- and right-hand sides of each equation. Results are summarized per chromosome in Supplementary Table 1, reporting the median error and the worst (maximum) block error. In brief, properties 2-4 are almost always satisfied under a tolerance of $10^{-3}$. Property 1 is more variable. To characterize whether these deviations are diffuse or localized, we additionally computed the relative Frobenius error. This error remained low overall, indicating that departures from property 1 are largely driven by a small number of problematic variants rather than systematic instability. Since we recommend interpreting a variant as reliable only when it is significant in both GWAS and *LDeconv*, the impact of these localized deviations should be limited.

#### Calibration of the LD pseudo-inversion method using simulations

The choice of the LD matrix pseudo-inversion method, as well as its associated parameters, can have substantial consequences for both inversion quality and the performance of the *LDeconv* method. We evaluated several pseudo-inversion approaches mentioned in the literature:

- Shrinkage, replacing the LD matrix $\mathbf{LD}$ with a weighted combination of $\mathbf{LD}$ and the identity matrix before inversion:

$$\mathbf{LD}^{-1}_{\mathbf{shrinkage}}(\delta) = [(1-\delta)\mathbf{LD} + \delta\mathbf{I}]^{-1}, \qquad \delta \in \{0.1,\ 0.5,\ 0.9\}$$

- Ridge inversion, adding a ridge parameter to the diagonal of the LD matrix before inversion:

$$\mathbf{LD}^{-1}_{\mathbf{ridge}}(\lambda) = (\mathbf{LD} + \lambda\mathbf{I})^{-1}, \qquad \lambda \in \{0.1,\ 1,\ 10\}$$

- Truncated singular value decomposition (TSVD), retaining the smallest number $k$ of components required to reach the retained variance threshold $\tau$:

$$\mathbf{LD}^{+}_{\mathbf{TSVD}}(\tau) = \mathbf{V}_k\mathbf{\Sigma}_k^{-1}\mathbf{U}_k^{\top}, \qquad \tau \in \{0.90,\ 0.95,\ 0.99,\ 0.999\}$$

- TSVD followed by ridge inversion, adding a ridge parameter to the retained singular values before inversion:

$$\mathbf{LD}^{+}_{\mathbf{TSVD+ridge}}(\tau,\lambda) = \mathbf{V}_k(\mathbf{\Sigma}_k + \lambda\mathbf{I}_k)^{-1}\mathbf{U}_k^{\top}, \qquad (\tau,\lambda) \in \{(0.95{,}1),\ (0.99{,}0.1),\ (0.99{,}1)\}$$

To limit computational costs, these benchmarks were performed on chromosome 22. Across 50 replicates for each of two representative simulation settings (heritability = 0.5 and polygenicity = 0.001, or heritability = 0.8 and polygenicity = 0.01, both with a sample size of 500,000), we measured the root mean square error (RMSE) between the observed and deconvolved Z-scores, separately for causal and non-causal variants, as well as the precision and recall for identifying causal variants at a significance threshold of $p < 5 \times 10^{-8}$. The results are presented in Supplementary Figures 1 and 2.

By prioritizing precision while accounting for recall, while also considering empirical performance on real data, we found that TSVD with a retained variance threshold of 0.99 provided the best overall performance for our summary-statistics deconvolution approach, *LDeconv*.

#### Simulations with varying sample sizes

We replicated the analyses presented in the Main Figure 2 using sample sizes of $100{,}000$ and $1{,}000{,}000$ and report the results in Supplementary Figures 6 and 7. As expected, the performance improved with larger sample size as the observed effects become more easily detectable, and decreased with smaller sample size.

#### Impact of ancestral divergence on the precision of *LDeconv*

To evaluate the cross-population portability of the UK Biobank (UKB)-derived inverse LD matrices, we simulated GWAS signals using LD structures from three distinct subsets of the 1000 Genomes Project (1000G): European (EUR), a global composite (ALL), and African (AFR) samples. As illustrated in Supplementary Figure 8, the precision of causal variant identification was highest when the GWAS LD structure closely aligned with the UKB reference panel ancestry (1000G EUR). However, a stepwise reduction in precision could be observed as the ancestral composition of the discovery cohort diverges, with the global population (1000G ALL) exhibiting moderate robustness and the African subset (1000G AFR) demonstrating a significant drop in precision. This degradation highlights the impact of distinct LD architectures and substantial differences in recombination rates, which prevent the truncated SVD from accurately isolating causal variants across diverged populations. These findings confirm that while *LDeconv* is resilient to minor cohort-specific variations, its reliability strictly depends on the use of an ancestrally compatible LD reference panel.

#### Sensitivity to LD matrix misspecification

To further evaluate the numerical stability of *LDeconv* against structural discrepancies in the Linkage Disequilibrium (LD) reference, we performed a sensitivity analysis by introducing controlled noise into the LD matrix prior to inversion. This noise was modeled through linear interpolation between the reference LD matrix ($\mathbf{LD}$) and its 180-degree rotated counterpart ($\mathbf{LD}rot180$), following the expression: $\mathbf{LD}noisy = a\mathbf{LD} + (1-a)\mathbf{LD}rot180$, where $a$ represents the interpolation weight.

We focused our evaluation on precision, defined as the proportion of true causal variants among those identified as significant. As shown in Supplementary Figure 9, *LDeconv* demonstrated robustness to minor matrix perturbations, maintaining stable precision levels for noise at or below 5% ($a \geq 0.95$). However, we observe a strict degradation in precision when the noise level exceeds this threshold ($a < 0.95$). These results indicate that while the truncated SVD framework provides a buffer against minor structural errors, the reliability of the deconvolved estimates depends on a reasonably accurate local LD structure.

#### Benchmark of the inversion time

Since *LDeconv* requires in-sample LD, the computational performance of inverting a new LD matrix must be evaluated. We benchmarked the inversion of chromosome 21, one of the smallest chromosomes, but one that contains comparatively large LD blocks. Runtime and maximum RAM usage were measured across three independent runs while parallelizing the computation across 1, 5, and 10 CPUs. We inverted each block sequentially, using Docker to restrict the process to the selected number of CPUs.

As shown in Supplementary Figure 10, a computational bottleneck appears to limit performance, such that increasing the number of CPUs does not reduce runtime. An alternative parallelization strategy, in which one block was assigned per CPU across multiple CPUs, did not improve runtime either. While additional CPUs may prove beneficial for larger LD blocks, we preferred to use comparatively smaller LD blocks combined with a sliding-window approach to maintain computational affordability. Nevertheless, the total runtime on a single CPU and the associated RAM usage remains reasonable, indicating that genome-wide LD inversion can be performed on standard computing hardware within a few days.
